# Reduced expression of fMRI subsequent memory effects with increasing severity across the Alzheimer’s disease risk spectrum

**DOI:** 10.1101/2023.09.11.23295362

**Authors:** Joram Soch, Anni Richter, Jasmin M. Kizilirmak, Hartmut Schütze, Slawek Altenstein, Peter Dechent, Klaus Fliessbach, Wenzel Glanz, Ana Lucia Herrera, Stefan Hetzer, Enise I. Incesoy, Ingo Kilimann, Okka Kimmich, Dominik Lammerding, Christoph Laske, Andrea Lohse, Falk Lüsebrink, Matthias H. Munk, Oliver Peters, Lukas Preis, Josef Priller, Ayda Rostamzadeh, Nina Roy-Kluth, Klaus Scheffler, Anja Schneider, Annika Spottke, Eike Jakob Spruth, Stefan Teipel, Jens Wiltfang, Frank Jessen, Emrah Düzel, Björn H. Schott

## Abstract

In functional magnetic resonance imaging (fMRI) studies, episodic memory is commonly investigated with the subsequent memory paradigm in which brain activity is recorded during encoding and analyzed as a function of subsequent remembering and forgetting. Impaired episodic memory is common in individuals with or at risk for Alzheimer’s disease (AD), but only few studies have reported subsequent memory effects in AD or its risk states like mild cognitive impairment (MCI). One reason for this might be that subsequent memory responses may be blunted in AD or MCI and thus less likely to manifest in fMRI signal differences. Here, we used Bayesian model selection of single-subject fMRI general linear models (GLMs) for a visual novelty and memory encoding experiment to compare the model performance of categorical and parametric subsequent memory models as well as memory-invariant models in a clinical cohort (N = 468) comprising healthy controls (HC) as well as individuals with subjective cognitive decline (SCD), MCI, and AD, plus healthy relatives of AD patients (AD-rel). We could replicate the previously reported superiority of parametric subsequent memory models over categorical models (Soch et al., 2021a) in the HC and also in the SCD and AD-rel groups. However, memory-invariant models outperformed any model assuming subsequent memory effects in the MCI and AD groups. In the AD group, we additionally found substantially lower model preference for models assuming novelty compared to models not differentiating between novel and familiar stimuli. Our results suggest that voxel-wise memory-related fMRI activity patterns in AD and also MCI should be interpreted with caution and point to the need for additional or alternative approaches to investigate memory function.

## 1. Introduction

Episodic memory, the ability to store, maintain and recall past singular events (Tulving, 1985), is highly vulnerable to impairment in dementia, and particularly in Alzheimer’s disease (AD), the most prevalent cause of dementia in old age (Livingston et al., 2017, 2020). As AD-related neuropathological changes precede clinically manifest dementia by several years (Jack et al., 2000; Chételat et al., 2005; Ledig et al., 2018), it is important to define pre-clinical stages of AD and risk states, which describe intermediate stages between (age-adjusted) normal cognitive functioning ability and manifest disease. Mild cognitive impairment (MCI), a measurable decline of cognitive function, but with preserved ability to perform activities of daily living (Petersen, 2016), is a widely recognized risk state for dementia due to AD, and, more recently, subjective cognitive decline (SCD), has been identified as a pre-MCI risk state, particularly when associated with worries about ones perceived cognitive deficit (Jessen et al., 2014, 2020). Both SCD and MCI do not *per se* constitute prodromal stages of AD, but they are associated with elevated risk to develop AD, particularly in individuals with the AD-typical findings in cerebrospinal fluid (CSF), that is, reduced levels of amyloid beta (Aβ1-42), and increased levels of total tau (tTau) and especially phosphorylated tau (pTau181) (Blennow et al. 2010; Olsson et al. 2016; Mattson-Carlgren et al., 2023).

In cognitive and clinical neuroscience, episodic memory is typically operationalized by various memory encoding and retrieval paradigms (Richardson-Klavehn & Bjork, 1988; Yonelinas, 2002), where both encoding (e.g., incidental vs. intentional) and retrieval task (e.g., recognition or free recall) can vary. An encoding task followed by a later memory test is frequently employed in neuroimaging studies of episodic memory, to infer on neural correlates of successful encoding (i.e., later memory) by comparing neural responses to remembered versus forgotten items (Brewer et al., 1998; Wagner et al., 1998; Kim, 2011; Maillet & Rajah, 2014). Neural differences related to encoding success are often referred to as subsequent memory effect (SME; also DM effect, for “difference due to memory”, see Düzel et al., 2011). Including both novel and previously familiarized stimuli in the encoding task allows to additionally infer on neurocognitive underpinnings of novelty processing (novel vs. familiar items; Henson et al., 2002), which show substantial, but not complete, overlap with the SME (Maass et al., 2014; Soch et al., 2021b).

Given the pronounced episodic memory deficits in Alzheimer’s disease and, to a lesser extent, also in MCI, applying the subsequent memory paradigm to people with Alzheimer’s risk states appears as a plausible approach to investigate the functional neuroanatomical correlates of AD-related memory impairment. Indeed, numerous functional magnetic resonance imaging (fMRI) studies of memory encoding and retrieval have been conducted in patients with early-stage AD or MCI (Gould et al., 2005; Kircher et al., 2007; Browndyke et al., 2013; Terry et al., 2015; Billette et al., 2022; Düzel et al., 2022) as well as in individuals with endogenous or exogenous risk factors for AD (Bookheimer et al., 2000; McDonough et al., 2020). Converging meta-analytic evidence suggests that individuals with AD or MCI exhibit reduced memory-related hippocampal activation and increased activation of medial parietal structures like the precuneus during encoding (Browndyke et al., 2013; Terry et al., 2015), and a similar pattern has been found to covary with increased risk for AD in clinically unaffected individuals (McDonough et al., 2020). It must be cautioned, though, that only few studies included in the aforementioned meta-analyses actually reported SMEs (Gould et al., 2005; Kircher et al., 2007; Trivedi et al., 2008), whereas others compared encoding to a low-level baseline or reported novelty effects (Browndyke et al., 2013; Billette et al., 2022).

While the reasons for not conducting a subsequent memory comparison between patients with AD or MCI and healthy controls were not typically reported, it seems plausible to assume that low memory performance and disintegration of memory networks might have resulted in a signal-to-noise ratio that is too low to compute meaningful subsequent memory contrasts in the clinical groups. Therefore, we aimed to employ an unbiased approach to assess the utility of subsequent memory models in comparison to memory-invariant novelty/familiarity models across the spectrum of AD risk. We have previously applied Bayesian model selection (BMS) to fMRI data acquired during an incidental visual episodic memory encoding task followed by a recognition memory task with a 5-point recognition-confidence rating scale (ranging from “item sure new” over “don’t know” to “item sure old”). We found that, in healthy young and older adults, SME models (taking encoding success into account) outperformed memory-invariant novelty models and, among SME models, (particularly non-linear) parametric models performed better than categorical models of the fMRI subsequent memory effect (Soch et al., 2021a).

To assess whether this pattern replicates for individuals with AD or at increased risk for AD, we applied the exact same approach to a large clinical cohort from the DZNE Longitudinal Cognitive Impairment and Dementia Study (DELCODE; Jessen et al., 2018), a memory-clinic-based multi-center study that focuses on individuals with SCD. Our sample consisted of healthy older control participants (HC, N = 128) and individuals with SCD (N = 199), MCI (N = 74), or early-stage Alzheimer’s disease (AD, N = 21), as well as first-degree relatives of patients with AD (AD-rel, N = 46). Importantly, all participants performed the exact same experimental paradigm as in the original study, were scanned with the same MRI protocol, and their data were analyzed using the same space of fMRI models as in the original study.

The goal of the present study was two-fold: First, we aimed to assess the replicability of the model preferences found in the original study in the sub-sample of healthy older adults (HCs). Second, we sought to identify differences in the preferences for fMRI episodic memory encoding models across the spectrum of AD risk states (HC → SCD → MCI → AD). We hypothesized that more severely affected individuals (AD and, to some extent, MCI) would exhibit less pronounced model preferences for novelty and particularly subsequent memory models, up to the point that, in AD patients, no model including an SME would outperform a memory-invariant model. Such a result would suggest that encoding-related fMRI signals in AD patients differentiate poorly, if at all, between subsequently remembered and forgotten items, and it would help to explain the previously described variability of between-group differences (McDonough et al., 2020) and the low proportion of studies reporting actual SMEs among the sizable number of fMRI studies on memory encoding in AD and also MCI (Terry et al., 2015; Nellessen et al., 2015).

## 2. Methods

### 2.1. Study cohort

In the present study, we applied a previously described methodology, validated in a cohort of healthy young and older adults (Soch et al., 2021a), to a sample of individuals with SCD, MCI, and early-stage AD as well as healthy controls and first-degree relatives of AD patients from the DZNE Longitudinal Cognitive Impairment and Dementia Study^1^ (DELCODE; Jessen et al., 2018), a multi-center study based in memory clinics collaborating with the German Center for Neurodegenerative Diseases (DZNE). All participants groups except HC and AD-rel were memory-clinic referrals and underwent clinical assessments, including a medical history interview, psychiatric and neurological examinations, neuropsychological testing, and routine MRI scans. Cognitive functioning was assessed using the Consortium to Establish a Registry for Alzheimer’s Disease (CERAD; Fillenbaum et al., 2008) neuropsychological test battery and the Mini Mental Status Examination (MMSE; Beyermann et al., 2013). Participants in the HC and AD-rel groups were recruited via newspaper advertisements.

The diagnostic assignment of participants to groups was as follows: Participants were assigned to HC, if they self-identified as cognitively healthy, passed a telephone screening for SCD, and their memory test performance was within 1.5 standard deviations (SD) of age-, gender-, and education-adjusted normal performance on *all* CERAD subtests. If participants expressed concerns regarding cognitive decline, a semi-structured interview with a physician at a memory clinic was conducted, and following the SCD-plus criteria (Jessen et al., 2014) and their CERAD performance, they were assigned to either SCD or MCI. Whereas MCI was assigned, if participants performed more than 1.5 SD below normal on the “recall word list” subtest, SCD was assigned, when they performed above this threshold. Participants with non-amnestic MCI were excluded from participation (Jessen et al., 2018), resulting in the MCI group including only individuals with amnestic MCI. MCI patients also needed to not meet the criteria for AD. In the AD group, only patients with >18 and <26 points in the MMSE were included (Beyermann et al., 2013).

Complete baseline data (i.e., data from the first study visit) was available for 844 subjects at the time of data analysis. From these subjects, we had to exclude (i) subjects without available diagnosis, (ii) subjects who did not perform the fMRI task, (iii) subjects whose logfiles from the fMRI experiment were missing, and (iv) subjects lacking one or more MRI data files necessary for our pre-processing pipeline (see below). This resulted in a final sample size of N = 468 subjects (HC: 128; SCD: 199; MCI: 74; AD: 21; AD-rel: 46) used for the analyses reported here. Demographic information for the different groups is given in Table 1.

**Table 1.** *Demographic information for participant groups*, along with multi-group comparisons (column “Statistics”) as well as pair-wise tests against healthy controls (rows “test vs. HC”). Statistical inference was based on one-way ANOVAs and two-sample t-tests (age), Kruskal-Wallis H-tests and Mann-Whitney U-tests (MMSE) as well as chi-square tests for independence (gender, site). Please note that neuropsychological testing results for the same cohort are reported in the accompanying second part of this paper (Soch et al., 2023b). Abbreviations: HC = healthy controls, SCD = subjective cognitive decline, MCI = mild cognitive impairment, AD = Alzheimer’s disease, AD-rel = AD relatives; N = sample size, yrs = years, m = male, f = female, MMSE = mini-mental state examination (Folstein et al., 1975; Jessen et al., 2018. Significance: * p < 0.05, ** Bonferroni-corrected for number of comparisons per variable (4).

|  | HC | SCD | MCI | AD | AD-rel | Statistics |
| --- | --- | --- | --- | --- | --- | --- |
| sample size | N = 128 | N = 199 | N = 74 | N = 21 | N = 46 | – |
| age range | 60-87 yrs | 59-85 yrs | 62-86 yrs | 60-80 yrs | 59-77 yrs | – |
| mean age | 69.27 ± 5.48 yrs | 70.36 ± 5.88 yrs | 72.98 ± 5.13 yrs | 72.56 ± 5.41 yrs | 65.91 ± 4.69 yrs | $F_{4,463} = 13.50$,<br>$p < 0.001$ |
| test vs. HC | – | $t_{325} = 0.89$, $p = 0.372$ | $t_{200} = 4.19$, $p < 0.001^{**}$ | $t_{147} = 2.19$, $p = 0.030^*$ | $t_{172} = -4.31$, $p < 0.001^{**}$ | |
| gender ratio<br>(male/female) | 48/80 m/f | 109/90 m/f | 35/39 m/f | 8/13 m/f | 18/28 m/f | $\chi^2_4 = 11.26$,<br>$p = 0.024$ |
| test vs. HC | – | $\chi^2_1 = 9.31$, $p = 0.002^{**}$ | $\chi^2_1 = 1.86$, $p = 0.173$ | $\chi^2_1 = 0.00$, $p = 0.958$ | $\chi^2_1 = 0.04$, $p = 0.845$ | |
| acquisition site<br>(8 centers) | 37 / 16 / 2 / 35 /<br>9 / 3 / 18 / 8 | 42 / 20 / 17 / 29 / 24 / 9 /<br>53 / 5 | 12 / 9 / 8 / 11 / 3 / 2 /<br>28 / 1 | 5 / 0 / 1 / 8 / 3 / 0 / 2 / 2 | 7 / 6 / 7 / 6 / 7 / 1 / 11 / 1 | $\chi^2_{28} = 60.19$,<br>$p < 0.001$ |
| test vs. HC | – | $\chi^2_7 = 26.78$, $p < 0.001^{**}$ | $\chi^2_7 = 29.31$, $p < 0.001^{**}$ | $\chi^2_7 = 6.67$, $p = 0.464$ | $\chi^2_7 = 23.18$, $p = 0.002^{**}$ | |
| MMSE total | 29.43 ± 0.87 | 29.17 ± 1.10 | 28.05 ± 1.56 | 24.52 ± 3.75 | 29.48 ± 0.89 | $\chi^2_4 = 107.43$,<br>$p < 0.001$ |
| test vs. HC | – | $z = -2.20$, $p = 0.028^*$ | $z = -7.24$, $p < 0.001^{**}$ | $z = -7.20$, $p < 0.001^{**}$ | $z = 0.46$, $p = 0.645$ | |

### 2.2. Comparison with original study

Apart from using a different study cohort, comprising five (HC, SCD, MCI, AD and AD relatives) rather than two (healthy young and older adults) groups of participants, the present study uses the exact same workflow and protocols for data acquisition and data analysis as the original study (see Supplementary Table S2). While data acquisition mostly took place before completion of this original study used as the reference here (Soch et al., 2021a), the complete data analysis was performed after its publication, following the approval of a detailed analysis protocol by the DELCODE steering committee, such that the present work can be considered effectively preregistered^2^.

### 2.3. Experimental paradigm

Participants performed an incidental memory task introduced by Düzel and colleagues (Düzel et al., 2011), which was slightly adapted as part of the DELCODE protocol (Düzel et al., 2018; Bainbridge et al., 2019), with the adapted version also used in the “Autonomy in Old Age” study (Soch et al., 2021a; Soch et al., 2021b; Richter et al., 2023). Subjects were presented with photographs of indoor and outdoor scenes, which were either novel to the participant at the time of presentation (44 indoor and 44 outdoor scenes) or were repetitions of two pre-familiarized “master” images (22 indoor and 22 outdoor trials). In a later retrieval session, participants were presented with all novel images from the encoding session, now considered “old” stimuli (88 images), as well images not previously seen by the participant, i.e., “new” stimuli (44 images). Participants were asked to provide a recognition-confidence rating for each image, using a five-point Likert scale ranging from “sure new” (1) over “don’t know” (3) up to “sure old” (5). For further details, see previous descriptions of the paradigm (Assmann et al., 2020; Soch et al., 2021a; Soch et al., 2021b; Richter et al., 2023). To help participants focus their attention on the stimuli, responses were given overtly and recorded by a trained experimenter. There was also no response deadline in the retrieval task, and the next stimulus was only shown after a response had been given.

### 2.4. MRI data acquisition

MRI data were acquired at eight different sites of the DZNE across Germany (see Table 1), using Siemens 3T MR tomographs. All sites followed the exact same MRI protocol implemented in the DELCODE study (Jessen et al., 2018; Düzel et al., 2018). Structural MRI included a T1-weighted MPRAGE image (voxel size = 1 x 1 x 1 mm) as well as phase and magnitude fieldmaps for later spatial artifact correction. Functional MRI consisted of 206 T2*-weighted echo-planar images (TR = 2.58 s, voxel size = 3.5 x 3.5 x 3.5 mm) measured during the encoding session of the memory task (09:01 min) as well as a resting-state session comprising 180 scans (same parameters) not used for the present study. For detailed scanning parameters, see previous descriptions of data acquisition (Soch et al., 2021a, 2021b).

### 2.5. MRI data processing

MRI data were analyzed with Statistical Parametric Mapping^3^, version 12, revision 7771 (SPM12 R7771; Wellcome Trust Center for Neuroimaging, University College London, London, UK). Preprocessing of the fMRI data included acquisition time correction (*slice timing*), head motion correction (*realignment*), correction of magnetic field inhomogeneities using the fieldmaps (*unwarping*), coregistration of the T1-weighted MPRAGE image to the mean functional image, segmentation of the coregistered MPRAGE image and subsequent normalization of unwarped EPIs into the MNI standard space (voxel size = 3 x 3 x 3 mm) using the transformation parameters obtained from segmentation, and finally, spatial smoothing of the functional images (FWHM = 6 mm).

Statistical analysis of the fMRI data was based on voxel-wise general linear models (GLMs) that included two onset regressors, one for novel images (novelty regressor) and one for the master images (master regressor), six head movement regressors obtained from realignment and a constant regressor representing the implicit baseline. This setup is referred to as the “baseline model” and was later varied (see Sections 2.6 and 2.7) in order to test specific hypothesis using Bayesian model inference.

### 2.6. Bayesian model selection

Bayesian model inference was performed via cross-validated Bayesian model selection (cvBMS; Soch et al., 2016), as implemented in the SPM toolbox for model assessment, comparison and selection (MACS; Soch & Allefeld, 2018). This technique proceeds by calculating the voxel-wise cross-validated log model evidence (cvLME) for each GLM, applied to each participant’s data. Then, the cvLME maps from all subjects and models are submitted to voxel-wise random-effects Bayesian model selection (RFX BMS; Stephan et al., 2008; Penny et al., 2009; Rosa et al., 2010). Whenever a particular analysis addresses a comparison of model families rather than individual models (see below), a cross-validated log family evidence (cvLFE) is calculated from the cvLMEs of all models belonging to a family, before entering cvLFEs into RFX BMS. Group-level analysis results in selected-model maps (SMMs) which indicate, for each voxel, the most frequently selected optimal model for describing the measured group fMRI data. For each model or family comparison, we report continuous SMMs which indicate, for the winning model, the likeliest frequency (LF) of this model, based on the posterior distribution over candidate models from RFX BMS. The LF can be interpreted as the proportion of subjects in the population for which this model best explains the measured fMRI data or, alternatively, as the probability that the measured fMRI data of an individual subject were generated by this model (Stephan et al., 2008; Soch et al., 2021a).

### 2.7. Overview of the model space

There are two groups of variations that were applied to the baseline model (see Table 3 and Figure 8 in the Appendix): First, there were variations of no interest, testing different ways of describing the encoding event as such, without regard for actual encoding success. Second, the baseline model was modified to include different variants of the subsequent memory effect.

Variations of no interest included (i) replacing event duration of 2.5 s (the actual trial duration; model family “GLMs_TD”) with an event duration of 0 s (assuming point events; model family “GLMs_PE”); (ii) collapsing novel and master images (model family “GLMs_00”) rather than modeling them as separate regressors (model family “GLMs_0”); and (iii) separating indoor and outdoor images (model family “GLMs_x2”) rather than collapsing them into a single regressor (model family “GLMs_x1”). For details regarding variations of no interest, see Soch et al. (2021a), Section 3.1.

Model modifications introducing a subsequent memory effect included (i) splitting novel images into 2, 3, or 5 categories based on the corresponding later memory responses (model family “GLMs_2” and models “GLM_3” and “GLM_5”); (ii) parametrically modulating the novelty regressor with theoretical (i.e. *a priori* defined) functions of the subsequent memory response (model family “GLMs_1t”); and (iii) parametrically modulating the novelty regressor with empirical (i.e. single-subject-data-derived) functions of the subsequent memory response (model family “GLMs_1e”).

Model “GLM_5” included five categorical regressors, one for each of the five response categories of the recognition-confidence scale (“sure new”, “probably new”, “don’t know”, “probably old”, “sure old”). Model “GLM_3” collapsed the “probably" and “sure” responses, resulting in three categorical regressors (“old”, “don’t know”, “new”). Model family “GLM_2” collapsed the five response options to two categorical regressors, where neutral responses were either considered forgotten (“GLM_2nf”) or remembered (“GLM_2nr”), or split between these two categories, according to responses frequencies (“GLM_2ns”).

The parametric model family “GLMs_1t” employed parametric modulations of the single novelty regressor with continuous functions of the participant’s responses on the recognition-confidence scale, namely either a linear transformation (“GLM_1t-l”), or an arcsine transformation (“GLM_1t-a”) or a sine transformation (“GLM_1t-s”), in order to either put a more weight on the “sure” responses than on the “probably” responses (arcsine) or weighting the “probably” responses more strongly than the linear model (sine).

The parametric model family “GLMs_1e” also employed parametric modulations of the single novelty regressor with a function the participant’s memory response, which was, in this case, not defined *a priori*, but instead based on individual response frequencies, using either conditional probability, inverse probability or a logistic regression model for modulator values (see Table 3). For further details about modelling the subsequent memory effect, see Soch et al. (2021a), Sections 3.2-3.3.

In total, these variations resulted in 19 first-level GLMs describing the fMRI data scanned during memory encoding (see Appendix, Table 3 and Figure 8). This model space is successively explored using model family and individual model comparisons (see Supplementary Table S1). For some of these analyses, models were grouped into families according to their abbreviations. For example, “GLMs_1” is the family of all parametric-modulator models, “GLMs_2” is the family of all categorical models with two regressors, “GLMs_12” is the family of all memory models with one or two regressors, etc.

### 2.8. Statistical analyses

In addition to group-level Bayesian model comparisons, classical analyses were performed on the single-subject extent of novelty and memory effects according to voxel-wise log Bayes factors (LBF). For this purpose, the number of voxels exceeding LBF > 3 (corresponding to a Bayes factor threshold of exp(3) ≈ 20) on either the comparison for novelty processing (comparing models separating novel and familiar items vs. models not doing so) or subsequent memory (comparing models with one or two memory regressors vs. the baseline model) was extracted from each subject’s LBF maps and used as the dependent variable.

These numbers were then subjected to an analysis of covariance (ANCOVA) with diagnostic group as categorical independent variable and additional factors and covariates of no interest (acquisition site, gender, age, years of education, and years of employment; see Figure 2 and Supplementary Table S3), followed by two-sample t-tests of each diagnostic group against healthy controls as well as Bayesian t-tests against healthy controls to quantify evidence for the hypothesis of no difference (see Table 2). Moreover, binary support vector classifications (SVC) were performed using both contrasts (novelty and memory) as features to assess separability of each diagnostic group from healthy controls (see Table 2).

**Table 2.** Bayesian model comparison for novelty processing and subsequent memory. Mean and standard deviations for number of voxels exceeding a log Bayes factor of 3 (approximately, a Bayes factor of 20) in Bayesian model comparisons testing for novelty and memory effects. Each participant group was tested against healthy controls (rows “vs. HC”) via two-sample t-tests and Bayesian t-tests. Additionally, support vector classifications were used to separate healthy controls from each other group based on number of supra-threshold voxels from both contrasts and using different log Bayes factor thresholds (corresponding to Bayes factors of 1, 3, 20 and 150). Abbreviations: t = t-statistic, p = p-value, BF_01_ = Bayes factor in favor of the null hypothesis, δ_med_ = posterior median effect size, BA = balanced accuracy, CI = 90% confidence interval. Significance: * p < 0.05, Bonferroni-corrected for ** number of tests per contrast (4) or *** number of tests and number of contrasts (4 x 2). This table summarizes results reported in Figure 2.

|  | HC | SCD | MCI | AD | AD-rel |
| --- | --- | --- | --- | --- | --- |
| *novelty processing (“GLMs_0” vs. “GLMs_00”)* |  |  |  |  |  |
| number of voxels with LBF > 3 | 5531.1 ± 3156.2 | 5127.6 ± 3103.0 | 4325.3 ± 3525.8 | 2391.1 ± 1560.5 | 6080.8 ± 3417.0 |
| two-sample t-test vs. HC | – | t<sub>325</sub> = 1.14, p = 0.255 | t<sub>200</sub> = 2.51, p = 0.013* | t<sub>147</sub> = 4.46, p < 0.001*** | t<sub>172</sub> = -0.99, p = 0.323 |
| Bayesian t-test vs. HC | – | BF<sub>01</sub> = 4.31, $\delta_{\text{med}}$ = 0.12 | BF<sub>01</sub> = 0.35, $\delta_{\text{med}}$ = 0.34 | BF<sub>01</sub> = 9.4·10<sup>-4</sup>, $\delta_{\text{med}}$ = 0.97 | BF<sub>01</sub> = 3.47, $\delta_{\text{med}}$ = -0.15 |
| *subsequent memory (“GLMs_12” vs. “GLM_TD_0x1”)* |  |  |  |  |  |
| number of voxels with LBF > 3 | 1605.1 ± 1454.6 | 1694.7 ± 1404.5 | 1045.1 ± 1001.5 | 800.1 ± 951.4 | 1636.8 ± 1656.0 |
| two-sample t-test vs. HC | – | t<sub>322</sub> = -0.55, p = 0.582 | t<sub>197</sub> = 2.93, p = 0.004*** | t<sub>143</sub> = 2.39, p = 0.018* | t<sub>169</sub> = -0.12, p = 0.903 |
| Bayesian t-test vs. HC | – | BF<sub>01</sub> = 6.88, $\delta_{\text{med}}$ = -0.06 | BF<sub>01</sub> = 0.12, $\delta_{\text{med}}$ = 0.40 | BF<sub>01</sub> = 0.36, $\delta_{\text{med}}$ = 0.50 | BF<sub>01</sub> = 5.38, $\delta_{\text{med}}$ = -0.02 |
| *novelty & memory (LBF thresholds 0.00, 1.10, 3.00, 5.01)* |  |  |  |  |  |
| SVM classification vs. HC | – | BA = 0.5085, CI=[0.45,0.56] | BA = 0.5693, CI=[0.50,0.64] | BA = 0.7157, CI=[0.58,0.83] | BA = 0.4422, CI=[0.35,0.53] |

ANCOVAs and two-sample t-tests were run in MATLAB R2018b using the functions “fitlm”, “anova” and “ttest2”. Bayesian t-tests were implemented in JASP 0.18.3 with a two-sided alternative hypothesis and the default Cauchy prior, reporting the Bayes factor in favor of the null hypothesis BF_10_ and the posterior median effect size δ_med_. For classification analyses, SVMs were calibrated with regularization hyperparameter C = 1 and using k = 10-fold cross-validation. To account for unequal sample sizes among participant groups, we repeatedly drew subsamples with a constant number of observations per class (N = sample size of smallest group). Classification accuracy and 90% confidence interval as measures of predictive performance were obtained as averages across all S = 1000 subsamples. These analyses were implemented using *Machine Learning for MATLAB* (https://github.com/JoramSoch/ML4ML).

## 3. Results

### 3.1. Participant groups differ by their behavioral response pattern

Behavioral response frequencies that were used as parametric modulators in the empirical parametric GLMs are shown in Figure 1. They included the conditional probability (i.e. the likelihood of a stimulus being old, given the subsequent memory response) and the inverse probability (i.e. the likelihood of a subsequent memory response, given the stimulus being old). Two patterns of variability across participant groups could be observed: First, “old” responses to old items (i.e. hits) became less frequent and “new” response to old items (i.e. misses) became more frequent when moving from HC towards AD (see Figure 1A). Second, the degree to which the subsequent memory response informs about an item being old diminished when moving from healthy controls towards AD patients (see Figure 1B). In both instances, healthy relatives of AD patients were qualitatively indistinguishable from healthy controls.^4^

**Figure 1.**
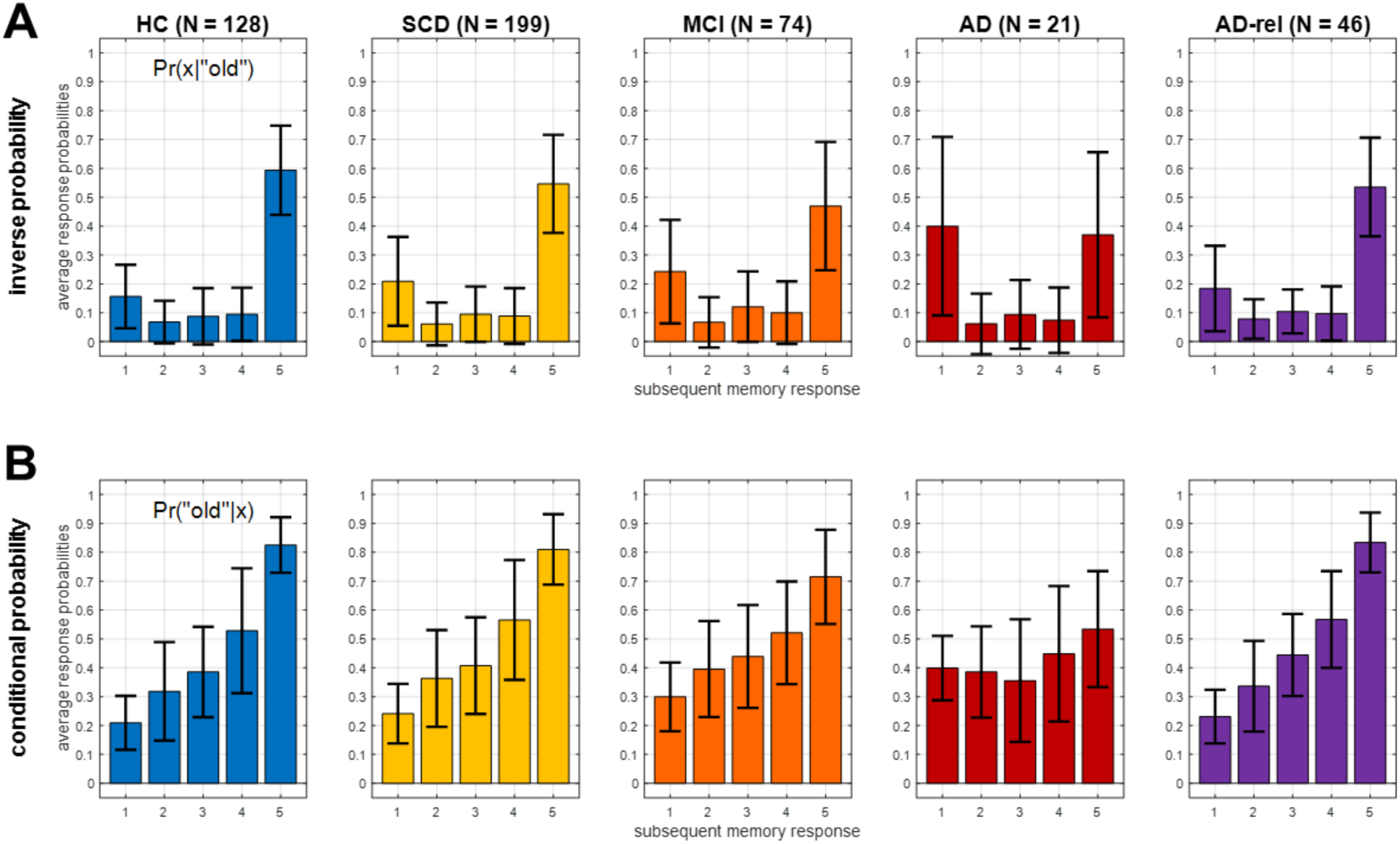
Behavioral response data across diagnostic groups. Empirically observed probabilities of **(A)** subsequent memory responses, given stimulus being old (“inverse probability”), and **(B)** stimulus being old, given a subsequent memory response (“conditional probability”). These probabilities were used as parametric modulators in the empirical parametric GLMs (see Table 3, model family “GLM_1e”). Error bars depict standard deviations (SD) across subjects. This figure corresponds to Figure 2B from the original publication.

**Figure 2.**
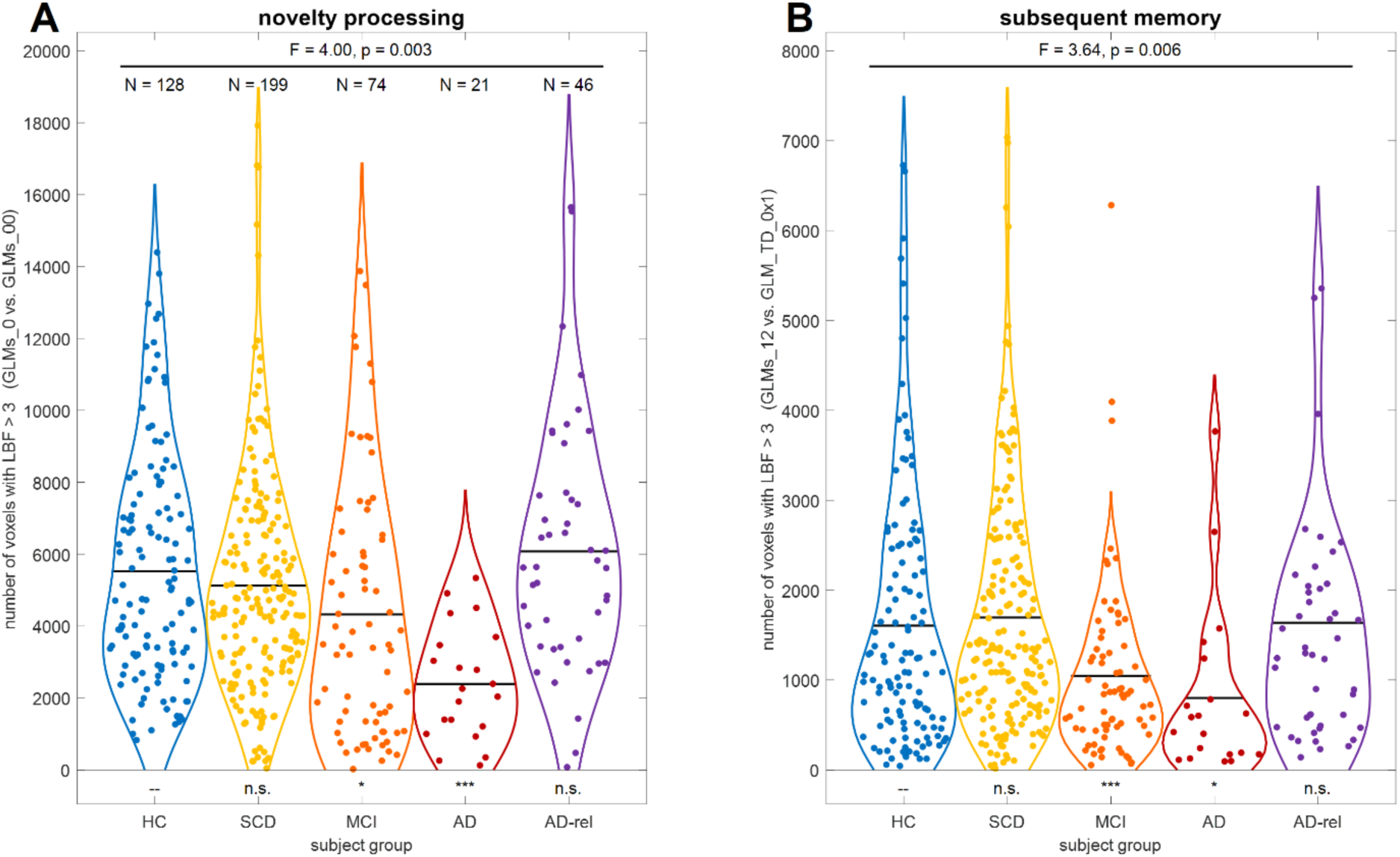
Bayesian model comparison for novelty processing and subsequent memory. Number of voxels exceeding a log Bayes factor of 3 (approximately, a Bayes factor of 20) in Bayesian model comparisons testing for **(A)** novelty processing (comparing models separating novel and familiar items vs. models not doing so) and **(B)** subsequent memory (comparing models with one or two memory regressors vs. the baseline model), for each subject from all five participant groups. Sample sizes are given in the left panel. Horizontal bars correspond to group-wise means. Statistics inside the panels correspond to the main effect of diagnostic group, controlled for gender, site, age, education and employment (F/p-value), as well as two-sample t-tests of each group against DELCODE healthy controls (significance markers). Abbreviations: HC = healthy controls, SCD = subjective cognitive decline, MCI = mild cognitive impairment, AD = Alzheimer’s disease, AD-rel = AD relatives. Significance: * p < 0.05, Bonferroni-corrected for ** number of tests per contrast (4) or *** number of tests and number of contrasts (4 x 2).

### 3.2. Variations of no interest are replicated in independent cohorts

Regarding modelling variations of no interest, i.e., modifications of the GLM unrelated to the subsequent memory effect, we could replicate all observations from the original study, albeit to a somewhat lesser degree in AD patients:

- First, the model family “GLMs_TD” was preferred throughout the gray matter in all subject groups (see Supplementary Figure S1), indicating that the actual trial duration of 2.5 s represents a better description of the measured neural signals than point events.
- Second, the model family “GLMs_0” was preferred in large clusters spanning temporal, occipital, and parietal cortical structures (see Figure 3A), indicating differential neural responses to novel vs. non-novel stimuli in these regions. Notably, this novelty effect was already diminished in AD patients (see Figure 3A, 4^th^ column).
- Finally, we observed that the model family “GLMs_x2” was preferred in selected portions of occipital cortex only (see Supplementary Figure S3), suggesting that the indoor-outdoor distinction was only important in a small subset of visual association cortices likely involved in scene processing. Since those regions were not the focus of our study, we omitted the indoor/outdoor distinction from the model, as in the original study with young and healthy HC (Soch et al., 2021a, p. 6).

**Figure 3.**
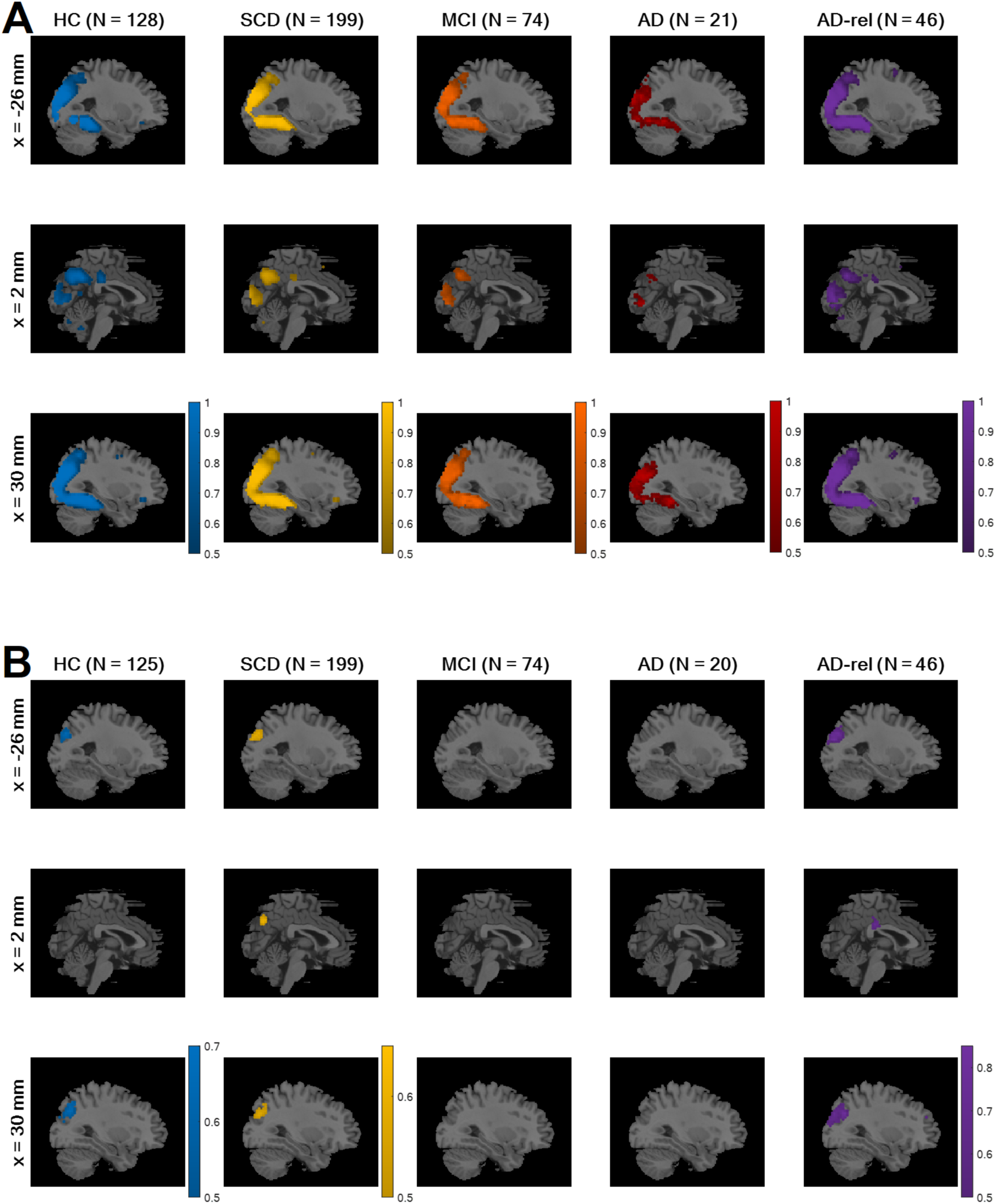
Modelling preferences for novelty processing and subsequent memory. Selected-model maps, showing **(A)** voxels with group-level preference in favor of the family of models separating novel and pre-familiarized images, against the family of models not considering novelty, and **(B)** voxels with group-level preference in favor of memory models, against the baseline GLM. Three sagittal slices (x-coordinates given at the left), roughly equal to those used in results display in the original study, are shown for each subject group (sample size given on top). Colored voxels display estimated group-level frequency **(A)** of the model family “GLMs_0” (novelty and master regressor), rather than the model family “GLMs_00” (both regressors collapsed), and **(B)** of the model family “GLMs_12” (one or two memory regressors), rather than the model “GLM_TD_0x1” (no memory effect). This figure corresponds to Figure S3B and Figure 3A from the original publication.

### 3.3. Subsequent memory effects decrease across the AD risk spectrum

Based on the outcomes described in Section 3.2., all following analyses were based on modifying a baseline model (i) using the actual trial length as event duration, (ii) modelling novel and master images separately, and (iii) collapsing indoor and outdoor images. First, we compared the family of models with either one parametric memory modulator or two categorical memory regressors (model family “GLMs_12”; i.e. models assuming a subsequent memory effect described using either one or two regressors) against the baseline GLM (model “GLM_TD_0x1”; i.e. a memory-invariant model).

While including an SME in the model improved the model fit in bilateral middle occipital gyrus (MOG), right temporo-parietal junction (TPJ) and the precuneus (PreCun) in the HC, SCD, and AD-rel groups (see Figure 3B; HC & AD-rel: no PreCun effect), memory-invariant models outperformed models considering subsequent memory performance in the MCI and AD groups (see Figure 3B, 3rd & 4th column).

To further substantiate the decline of memory – and, to some extent, novelty – effects across the AD risk spectrum, we performed a one-way ANOVA on the number of voxels with the respective model preferences, using diagnostic group as the between-subject factor. To this end, single subjects’ log Bayes factor (LBF) maps from model comparisons testing for effects of novelty processing (“GLMs_0” vs. “GLMs_00”; see Figure 2A) and subsequent memory (“GLMs_12” vs. “GLM_TD_0x1”; see Figure 2B), respectively, were generated, and the number of voxels exceeding LBF > 3 (corresponding to a Bayes factor threshold of exp(3) ≈ 20) was extracted as the dependent variable.

For both contrasts, there was a main effect of diagnostic group (novelty: F_4,446_ = 4.00, p = 0.003; memory: F_4,442_ = 3.64, p = 0.006) when additionally controlling for gender, site, age, educational years and employment years (see Supplementary Table S3 for detailed results), supported by significant differences of the MCI and AD, but not the SCD and AD-rel groups, from healthy controls. When using either a more liberal threshold (LBF > 1, corresponding to BF ≈ 3) or a more conservative threshold (LBF > 5, corresponding to BF ≈ 150), numbers of voxels were expectedly different, but the results were qualitatively identical in terms of observed effects and ranking of the groups.

Bayesian t-tests supported evidence for the null hypothesis (i.e., no group difference) when comparing HC against the SCD and AD-rel groups (all BF_01_ > 3.47), but not when comparing HC against the MCI and AD groups (all BF_01_ < 0.36; see Table 2). Furthermore, the number of supra-threshold voxels differentiated the AD and MCI groups from healthy controls when using SVM classification (see Table 2, last row).

### 3.4. Number of regressors effect increases across diagnostic groups

Among the GLMs modeling subsequent memory, we additionally tested for the influence of the number of regressors used to model the SME, which increases from the parametric memory models (1 parametric modulator per model) to the categorical memory models (2, 3 or 5 regressors; see Table 3). To this end, we calculated the LFE for each of these model families and subtracted the LME of the baseline GLM (0 memory regressors) to compute LBF maps in favor of memory models against a no-memory model. The rationale behind this was that some models assuming a memory effect might be too complex, thus performing even worse than memory-invariant models (see Soch et al., 2021a, Fig. 3B).

Note that the categorical model with five memory regressors (“GLM_5”) could only be estimated when each of the five behavioral response options occurred at least once. Therefore, these analyses were based on a subset of the participants (total N = 248; for N by group, see Figure 4). This procedure led to a very small N for the AD group, making the results for this group potentially less generalizable.

**Figure 4.**
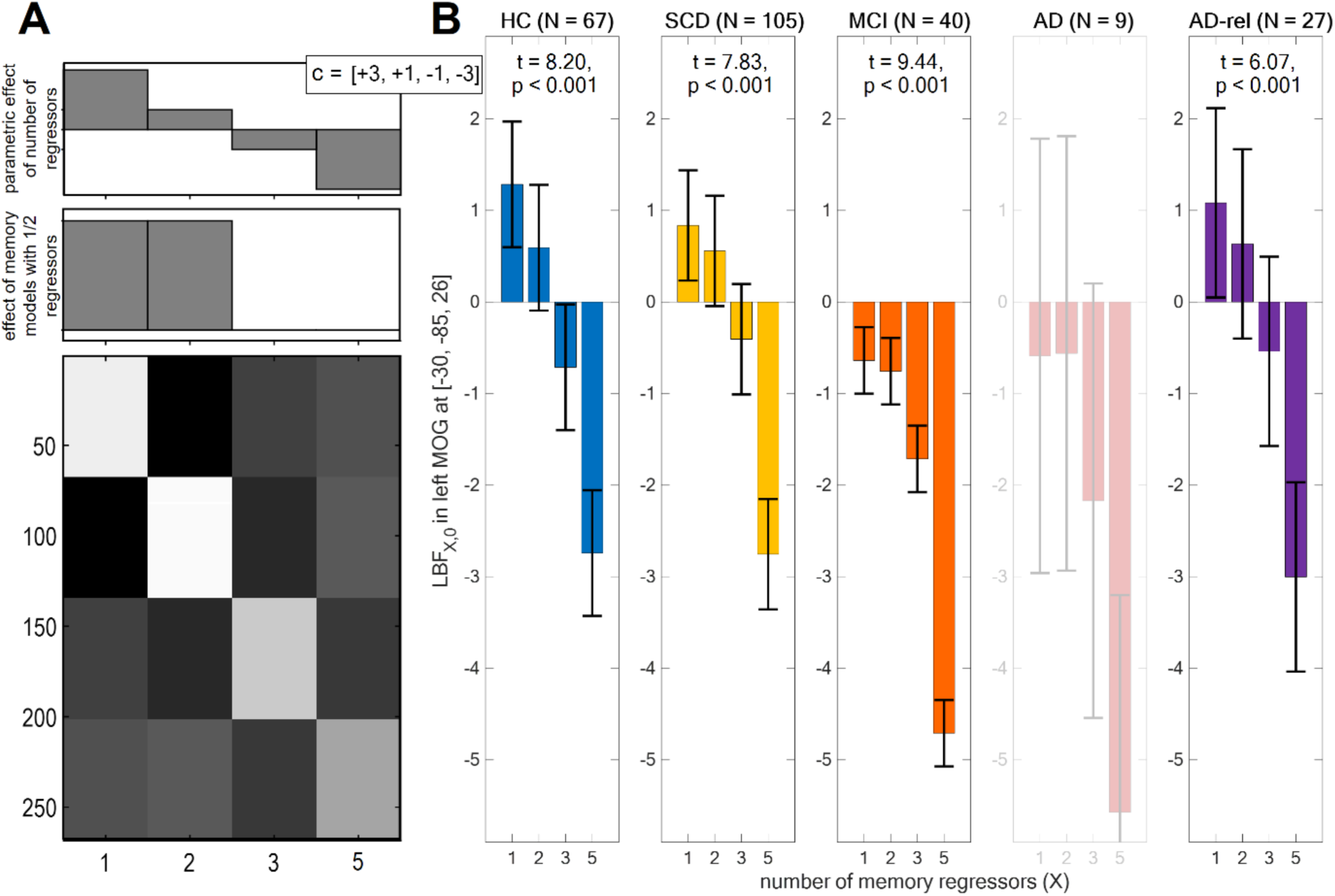
Influence of number of regressors on Bayesian model quality. **(A)** Design matrix of a second-level GLM in which log Bayes factor (LBF) maps comparing models with 1, 2, 3 or 5 memory regressors against the baseline GLM were submitted to a one-way ANOVA with dependencies between levels (here: number of regressors). A conjunction contrast of a significant memory effect (lower contrast) and a significant linear decrease of LBF with number of regressors (upper contrast) was used to identify coordinates of a global maximum in the original study. **(B)** Average LBFs from these coordinates in each group, along with t-statistics from the contrast testing for linear decrease of LBF with number of regressors. Bar plots depict contrasts of parameter estimates of the group-level model; error bars denote 90% confidence intervals (computed using SPM12). Note that this analysis could only be run for a subset of the participants, namely all those subjects that used the full range of behavioral responses, such that “GLM_5” could be fitted, hence the lower sample sizes in comparison to the other analyses (cf. Table 1). Bar plots are shaded and statistics are not reported for sample sizes smaller than 10. This figure corresponds to Figure 3B from the original publication.

The LBF maps were subjected to a one-way ANOVA model with the four-level within-subject factor number of regressors (see Figure 4A). There was a main effect of number of regressors throughout the whole brain (p < 0.05, FWE-corrected; results not shown). By performing a conjunction analysis between (i) a contrast of “GLMs_1” and “GLMs_2” against baseline and (ii) a t-contrast linearly decreasing with number of regressors (see Figure 4B), a global maximum was identified in the original study (see Soch et al., 2021a, Figure 3B). From the coordinates of that global maximum ([x, y, z] = [-30, -85, 26]; MNI coordinates in mm), LBFs were extracted to calculate parameter estimates, standard deviations and statistics for the linear contrast (see Figure 4B). These showed that GLMs with one or two memory regressors outperformed the memory-invariant model in the HC, SCD, and AD-rel groups, while they performed equally or even worse than the memory-invariant baseline GLM in the MCI and AD groups (see Figure 4B).

### 3.5. Parametric outperform categorical models in memory-related brain structures

When treating GLMs with one parametric modulator describing subsequent memory (“GLMs_1”) and categorical GLMs using two regressors for remembered vs. forgotten items (“GLMs_2”) as model families (i.e., calculating voxel-wise cvLFEs and comparing the two families via group-level cvBMS), we observed a preference for parametric GLMs throughout the memory network (see Figure 5A), especially in regions that also showed a novelty effect (cf. Figure 3A). The overall preference for parametric models was present in all diagnostic groups and extended to almost all voxels in the MCI and AD groups (see Figure 5A).

**Figure 5.**
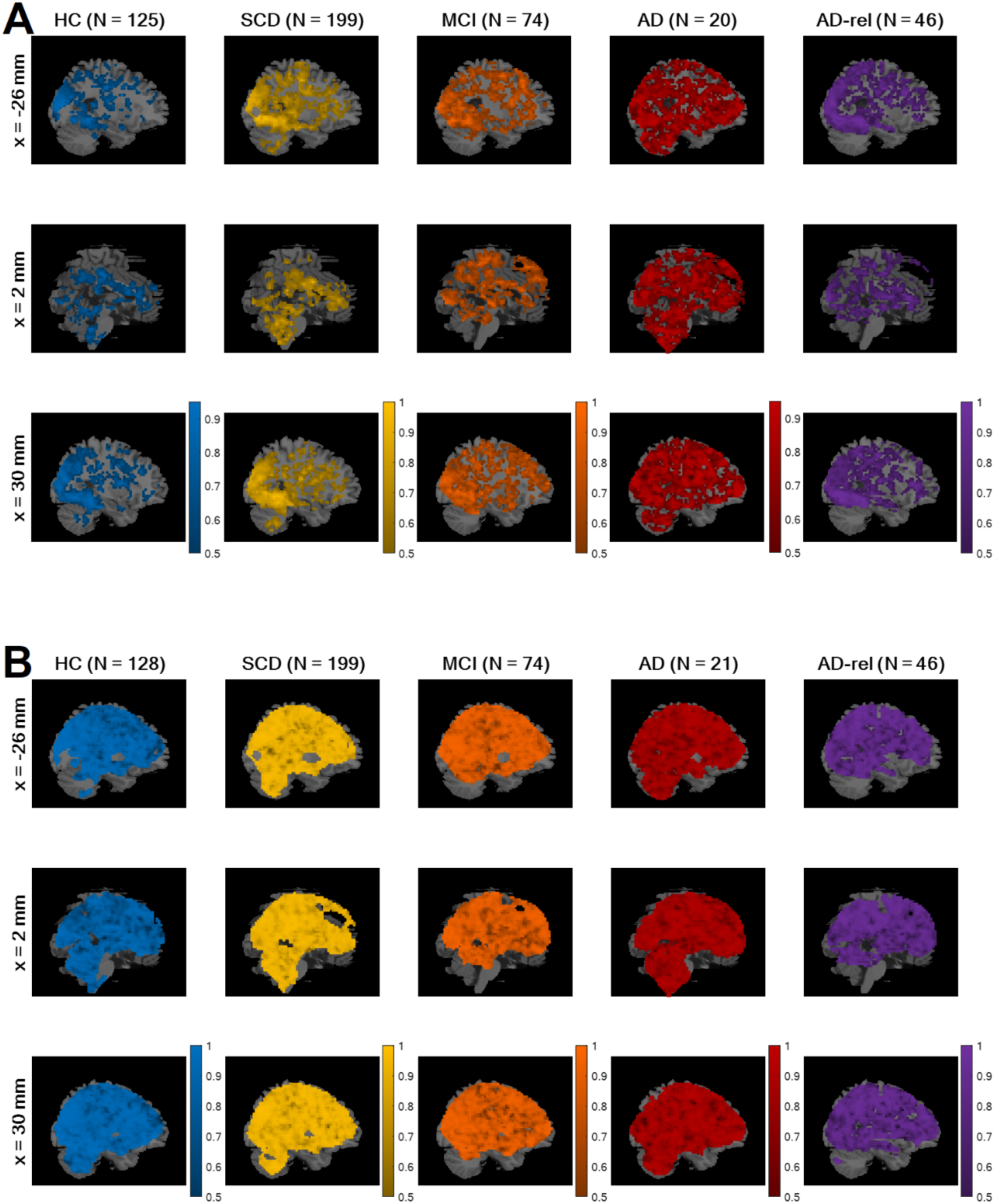
Bayesian comparison of memory model families. Selected-model maps, showing **(A)** voxels with group-level preference in favor of parametric models of the subsequent memory effect, using parametric modulators, against categorical models, separating response options, and **(B)** voxels with group-level preference in favor of empirical parametric models, using data-driven transformations, against theoretical models, using *a priori* defined transformations. The layout of the figure follows that of Figure 3. Colored voxels display estimated group-level frequency **(A)** of the model family “GLMs_1” (one memory regressor), rather than the model family “GLMs_2” (two memory regressors), and **(B)** of the model family “GLMs_1e” (empirical modulators), rather than the model family “GLMs_1t” (theoretical modulators). This figure corresponds to Figure 4 from the original publication.

Within the family of parametric memory models, we additionally compared theoretical GLMs (“GLMs_1t”) to empirical GLMs (“GLMs_1e”). Comparing these two sub-families via group-level cvBMS, we found an almost whole-brain preference for the empirical GLMs (see Figure 5B). This observation is in accordance with the original study with HC only (Soch et al., 2021a, Section 4.3) and was independent of disease status (see Figure 5B).

### 3.6. Model preferences within model families are replicated

Following the observation that models with one or two memory regressors outperform the memory-invariant model in large portions of the temporo-parieto-occipital memory network (see Figure 3B), we aimed to identify the optimal models within these different families. Within all sub-families of the memory models, we observed clear model preferences, consistent with observations in the original study (Soch et al., 2021a, Section 4.4):

- Among the two-regressor categorical GLMs, there was a clear preference for the model categorizing images with later neutral responses (response “3”) as forgotten items (“GLM_2-nf”), rather than either categorizing them as remembered items or randomly sampling neutral images as remembered or forgotten (see Figure 6A).
- Among the GLMs with theoretically based parametric modulators calculated, there was a clear preference for the model using an arcsine transformation of subsequent memory reports (“GLM_1t-a”) – which puts a higher weight on definitely remembered and forgotten items (responses “5” and “1”) –, rather than either a linear or a sine-transformed subsequent memory report (see Figure 6B). However, this preference was weaker in the AD group, possibly due a general deterioration of memory effects, in addition to a probably larger variance due to the relatively small size included individuals (N = 21).
- Within the GLMs with parametric modulators estimated from memory responses separately for each single subject, there was a clear preference for the model using the probability of “old” item given memory response as PM (“GLM_1e-ip”) over either employing the probability of memory response given “old” item as PM or estimating the conditional probability via a logistic regression model (see Figure 6C).

**Figure 6.**
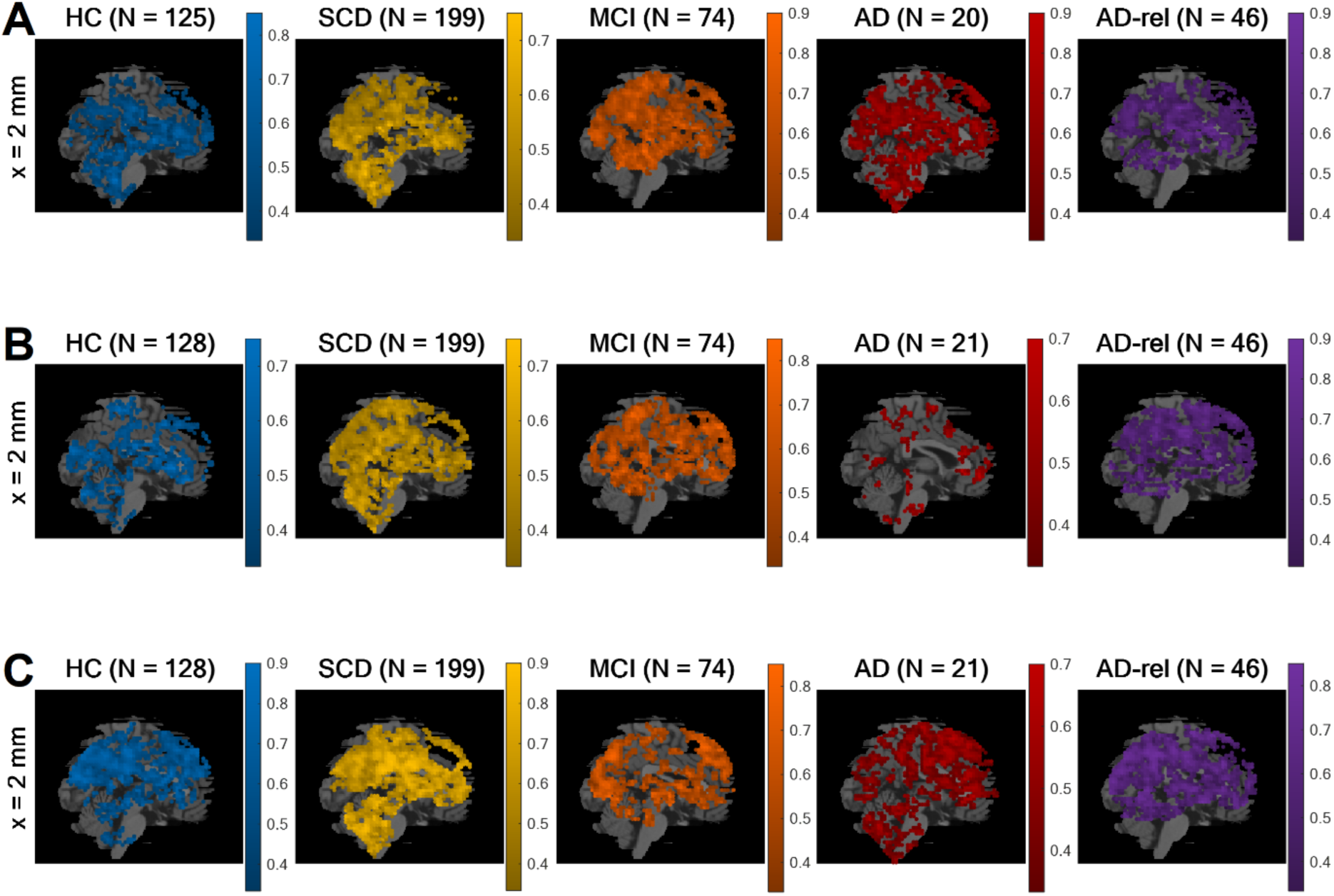
Winning models within memory model families. Selected-model maps in favor of **(A)** the GLM treating neutral images as forgotten items (“GLM_2-nf”), compared to the other two-regressor models (“GLM_2-nr”, “GLM_2-ns”), **(B)** the GLM using an arcsine-transformed parametric modulator (“GLM_1t-a”), compared to the other theoretical-parametric models (“GLM_1t-l”, “GLM_1t-s”), and **(C)** the GLM using an inverse probability parametric modulator (“GLM_1e-ip”), compared to the other empirical-parametric models (“GLM_1e-cp”, “GLM_1e-lr”). The layout of the figure follows that of Figure 3. Colored voxels display estimated group-level model frequencies. Due to clear model preferences, only one (the most medial) slice is shown for each comparison. This figure corresponds to Figure 5 from the original publication.

### 3.7. Novelty and memory parameter estimates reflect model preferences

Finally, in addition to the group-level Bayesian model selection – which informs us about the relative quality of different GLMs (e.g., parametric vs. categorical models) in explaining the measured BOLD signals –, we also performed group-level Frequentist statistical tests to probe statistically significant effects of task manipulations (novelty processing and subsequent memory) within each diagnostic group.

Specifically, we statistically tested for significantly positive or negative effects on (i) the novelty contrast from the GLM with arcsine-transformed PM (“GLM_1t-a”)^5^, (ii) the memory regressor from the parametric GLM with arcsine-transformed PM and (iii) from the parametric GLM with inverse probability PM as well as (iv) the memory contrast from the categorical GLM categorizing neutral responses as forgotten. All analyses were performed using F-contrasts in SPM, and a stringent family-wise error (FWE) correction at voxel level was applied (FWE, p < 0.05, k = 10). We observed two general patterns:

- First, the voxels showing statistically significant effects in a particular fMRI contrast showed a large overlap with those exhibiting model preferences in the respective model comparison. This pattern was found for both, novelty processing (cf. Figure 7A vs. Figure 3A) and subsequent memory (cf. Figure 7B vs. Figure 3B), and statistical significances are generally a bit stronger than model preferences (cf. Figure 7 vs. Figure 3).
- Second, there was a decline of novelty and memory effects across the AD risk spectrum, with (i) prototypical memory network activations in the HC, SCD, and AD-rel groups, (ii) reduced novelty effects and largely absent memory effects in individuals with MCI, and (iii) almost non-identifiable effects of both novelty and subsequent memory in AD patients (see Figure 7 and Supplementary Figures S10-S13).

**Figure 7.**
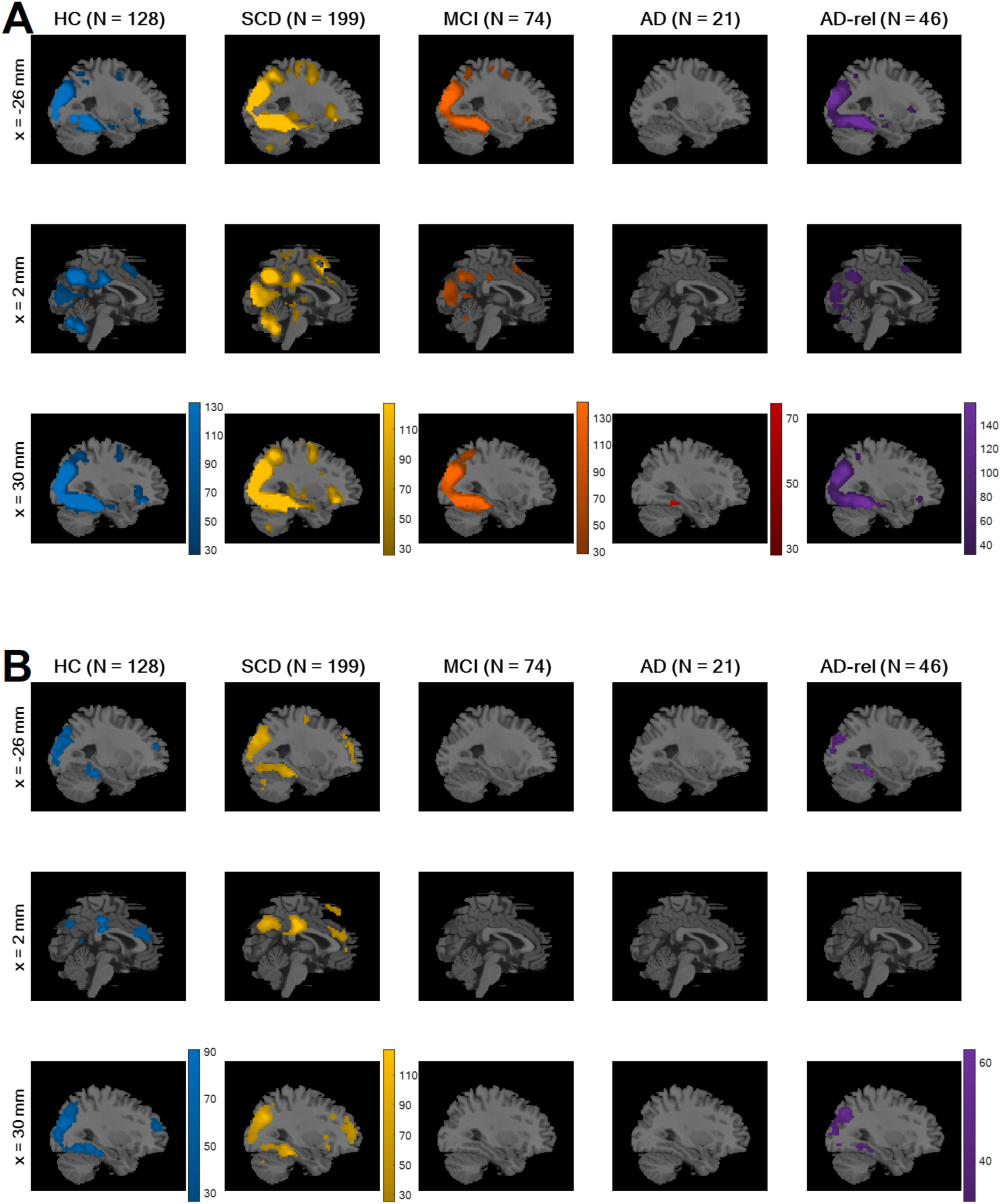
Novelty and memory effects from winning GLM. On the second level, a one-sample t-test was run across parameter estimates obtained from **(A)** the novelty contrast (novel vs. master images) and (B) memory contrast (parametric modulator) of the GLM using the arcsine-transformed PM. In SPM, statistical inference was corrected for multiple comparisons (FWE, p < 0.05, k = 10). Colored voxels display F-statistics indicating **(A)** significant (positive or negative) differences between novel and master images and **(B)** significant (non-zero) effects of the transformed memory response, on average across subjects from the respective participant group. This figure corresponds to Figure 7A and 7B from the original publication.

When reporting cluster-level-corrected instead of whole-brain FWE-corrected results for these analyses, one additionally observes mild effects of novelty processing in AD patients and mild effects of subsequent memory in MCI patients, but no memory effects for individuals with AD (see Supplementary Figure S14).

## 4. Discussion

In this study, we have applied cvBMS to fMRI data obtained during a visual novelty and memory encoding paradigm in older healthy controls (HC) and in individuals with AD or at an increased risk state (SCD, MCI, AD-rel). While we were able to replicate earlier findings regarding a superiority of parametric subsequent memory fMRI models (Soch et al., 2021a) in HC as well as in the SCD and AD-rel groups, we found that memory-invariant models largely outperformed subsequent memory models in individuals with MCI and AD and that manifest AD was associated with an additionally diminished novelty response.

### 4.1. Utility of parametric subsequent memory models in healthy older adults

We have previously demonstrated that subsequent memory models with one or two memory regressors are superior to memory-invariant models in healthy young and older adults and that, among the subsequent memory models, parametric models outperform categorical models (Soch et al., 2021a). In the present study, we were able to largely replicate this pattern of model preferences in the group of older healthy controls and also in individuals with SCD and in healthy relatives of AD patients. Assuming a novelty effect (i.e., a difference between novel and pre-familiarized *master* images) improves model quality in an extensive network including parietal, occipital and temporal cortices (hippocampus, parahippocampal and middle occipital gyri, MOG) as well as parts of the default mode network (precuneus, temporo-parietal junction, TPJ; see Figure 3A). Parts of this network further exhibited improved model quality when assuming a subsequent memory effect (see Figure 3B), particularly when employing a parametric subsequent memory model (see Figure 5A). As in our original study, BMS favored the model using an arcsine-transformed memory regressor among the theoretical parametric models (see Figure 6B), and the model using the inverse probability among the empirical models, (see Figure 6C).

As model family selection favored empirical over theoretical models, one might conclude that the model using the inverse probability would be the best-fitting model. However, it should be noted that a direct comparison of the two models in the original study yielded inconclusive results (Soch et al., 2021a). Furthermore, in all diagnostic groups, there were participants with a high number of (high-confidence) misses (see Figure 1). In such a situation, high-confidence hits and misses would both contribute to the “hits”, whereas items with low-confidence judgments would contribute to the “misses” of a regressor based on the inverse probability. This would rather reflect a participant’s response confidence than actual memory performance and thus constitute, at best, a questionable index of subsequent memory, despite providing a good model fit. Furthermore, different participants’ parametric modulators also operate at different scales which limits across-subject interpretability of their parameter estimates. High variability of response patterns among study participants would result in potentially large differences of the inverse probability regressor across subjects and possibly diagnostic groups, making group-level analyses difficult to interpret. We therefore recommend using the arcsine-transformed regressor that, like the inverse probability regressor, puts higher weight on high-confidence versus low-confidence hits which typically show more robust subsequent memory effects (Rugg et al., 2015; Hayes et al., 2017).

### 4.2. Decline of subsequent memory and novelty responses across the AD risk spectrum

Across the AD risk spectrum, we generally observed a progressive deterioration of memory model quality, with the effects of subsequent memory seen in HCs being largely preserved in the SCD and AD-rel groups, but practically absent in the MCI and AD groups (see Figures 2B/6B and Supplementary Figures S11-S13). Considering the rarity of studies reporting an actual subsequent memory effect rather than an encoding vs. baseline comparison (often a novelty effect) in patients with AD or MCI (Browndyke et al., 2013; Billette et al., 2022), our results point to the inherent difficulty of measuring a robust successful encoding response in these populations. In the group of AD patients, the absence of a robust fMRI effect (see Figure 7) was mirrored by a disrupted relationship between subsequent memory report (1-5) and item type (old vs. new) at the behavioral level (see Figure 1). This suggests that, with reduced behavioral accuracy, the predictive value of neural signals with respect to later remembering and forgetting also drops, such that fMRI responses do not covary with subsequent memory reports when the latter most likely reflect mere guessing, at least to a considerable proportion. On the other hand, the declining subsequent memory effect across the AD spectrum could also be interpreted as indicative of the memory decline, with a potential diagnostic or prognostic utility (Soch et al., 2024).

Despite showing a preserved, albeit diminished, relationship between subsequent memory report and item type (see Figure 1), individuals with MCI still exhibited, similarly to AD patients, a preference for memory-invariant models over subsequent memory models (see e.g. Figures 2B and 3B). This may be best explained by the notion that individuals with MCI, by definition, show clinically measurable deficits in memory performance (Petersen et al., 1999), and are thus likely to also exhibit higher guessing rates and thereby a decreased signal-to-noise ratio in fMRI subsequent memory models. On the other hand, not all individuals diagnosed with MCI eventually convert to AD (Grundman, 2004) which might be of importance when deriving putative biomarkers from fMRI data (Soch et al., 2023b).

Effects of novelty processing, on the other hand, were preserved, albeit at a reduced level, in individuals with MCI, but largely absent in patients with manifest AD (see Figures 2A and 6A). As the expression of a novelty effect requires prior successful familiarization of the baseline images (here called “master” images), we suggest that, at the MCI stage, encoding can still take place to some degree, for example with repeated presentation as was done with the master images. In the AD patients, on the other hand, familiarization might have been less effective, resulting in a poorer ability to distinguish novel from pre-familiarized images.

Despite the largely absent subsequent memory effect in the MCI and AD groups, the preference for parametric over categorical models of the subsequent memory effect was also evident in the MCI and AD groups and possibly even more pronounced than in the HC and SCD groups (see Figure 5A). In our view, the most parsimonious explanation for this observation is that, due to a pronounced reduction or even absence of the memory effect in the MCI and AD groups, BMS will inherently favor the parametric models due to their lower complexity.

The same pattern as in healthy older controls was, at least qualitatively, also observed in individuals with SCD and in healthy older relatives of patients with AD (see Figures 2, 3, 4, 6). Compatibly, individuals with SCD and healthy relatives also showed behavioral memory performance and fMRI signals comparable to that of healthy controls (see Figure 1 and 6). This points out the need to stratify SCD into those with subjective complaints and normal performance vs. those with subjective complaints and sub-normal performance (Koppara et al., 2015), possibly based on amyloid pathology (Jessen et al., 2022; Soch et al., 2024). Like the SCD group, healthy relatives often showed model preferences and activity patterns qualitatively identical to those of HCs (see e.g., Figures 2, 3 and 6), consistent with AD relatives in the DELCODE study not significantly differing from HCs in terms of MMSE total (see Table 1), NPT global score, the PACC5 neuropsychological composite score, or ApoE genotype (see Soch et al., 2023b). Thus, the overall preserved patterns of model preference in the SCD and AD-rel groups suggest that moderately increased clinical (SCD) or genetic (AD-rel) risk for AD is not *per se* associated with a disruption of functional memory network integrity.

### 4.3. Comparison with previous studies of memory encoding in AD and MCI

At first sight, our results seem to be at odds with previous studies reporting encoding-related fMRI activation differences between healthy controls and individuals with AD or MCI (for meta-analyses, see Browndyke et al., 2013; Nellessen et al., 2015; Terry et al., 2015). However, it must be noted that most studies contributing to those meta-analyses did not report subsequent memory contrasts in a strict sense, but contrasts comparing encoding against a low-level baseline or novelty contrasts (for exceptions, see Gould et al., 2005; Heun et al., 2007; Kircher et al., 2007; Trivedi et al., 2008). The broad inclusion of different encoding contrasts may explain the conflicting results of those meta-analyses with respect to encoding-related hippocampal activations (Terry et al., 2015: HC > AD; Nellessen et al., 2015: MCI > HC). A common finding in all three meta-analyses was, on the other hand, the relatively increased encoding-related activation of DMN structures, particularly precuneus.

The precuneus typically shows deactivations during successful encoding (Kim et al., 2011), which are attenuated in older adults (Maillet and Rajah, 2014; Kizilirmak et al., 2023; Schott et al., 2023). Deactivations of the precuneus are not specific to successful encoding, but have also been observed during novelty processing (Schott et al., 2023) and are more pronounced in individuals with SCD and MCI compared to HC (Billette et al., 2022). With respect to the present results, it must be noted that, at a more liberal threshold (p < 0.05, FWE-corrected at cluster level), we did observe reduced DMN deactivations in the MCI group during both, novelty processing and successful encoding (see Supplementary Figure S14).

It must be noted that our results with respect to the reduced expression of fMRI subsequent memory effects in AD and, to a lesser extent, also in MCI may not necessarily apply to other, for example, electrophysiological modalities like event-related potential (ERPs) to the same extent. ERP studies of successful encoding typically show frontal and centro-parietal positive deflections for subsequently remembered compared to subsequently forgotten stimuli (Paller et al., 1987; Fernández et al., 1998; Schott et al., 2002; Otten et al., 2007). While the importance of a sustained, positive potential (also referred to as “P600”) for successful encoding has been noted in the context of AD and MCI (Jackson and Snyder, 2008), a reduced P600 amplitude has thus far only been reported in the context of word repetition (Olichney et al., 2006). As such, it may correspond to the reduced or absent late positive component (LPC) in AD (Tendolkar et al., 1999), which likely reflects context-rich, recollection-based *retrieval* and depends on the integrity of the hippocampus (Düzel et al., 2001). Notably, the most recent available comprehensive review of cognitive ERP studies in AD and MCI (Paitel et al., 2021) did not include any studies of the subsequent memory effect. Therefore, while studies of repetition and retrieval point to abnormal memory-related ERPs in AD and MCI, specific alterations related to successful encoding are yet subject to future research.

### 4.4. Limitations and directions for future research

The primary benefit of this investigation lies in the use of cvBMS as an objective and unbiased procedure for voxel-wise fMRI model selection which accounts for both, model accuracy and model complexity, and allows for non-nested model comparison (unlike statistical significance tests on additional regressors; see Soch et al., 2016). A key limitation in this approach, however, is that cvBMS only provides information about the model quality without allowing for direct inferences on the sign or magnitude of a given regressor or contrast. For example, the preference for novelty or subsequent memory models within the default mode network (DMN) in a memory-impaired older person might originate from the prototypical encoding-related deactivations observed in healthy young and also cognitively unimpaired older individuals (Kim, 2011; Maillet & Rajah, 2014; Kizilirmak et al., 2023), which are associated with memory performance (Schott et al., 2023) and attenuated in individuals with memory impairment (Düzel et al., 2011; Maillet & Rajah, 2014; Billette et al., 2022) . Moreover, it cannot be excluded that cvBMS shows model preferences in voxels outside the networks of interest, even in white matter. This is simply a consequence of the fact that, in case of poor model fit, the most parsimonious model will be preferred. Therefore, one must cautiously examine the preferred models with respect to the plausibility of the voxels or clusters in which a model comes out as preferred model from cvBMS and, furthermore, complement cvBMS results by GLM-based analyses.

A limitation common to most fMRI studies of memory function in in AD is the low number of participants in the most severely affected AD group. While the number of 21 participants in this study was at the upper end compared to previous studies (see meta-analysis by Terry et al., 2015), it was nevertheless below the numbers desirable to obtain reproducible results (Button et al., 2013; Turner et al., 2018). Sample size was even smaller in the comparison of all models, which included only 9 individuals from the AD group, and we therefore refrained from interpreting the results of that group (see Figure 4). In addition to sample size, within-group heterogeneity in the clinical groups may potentially contribute to a lower signal-to-noise ratio and thus reduced expression of novelty-related and memory-related fMRI activation patterns. While we aimed to reduce such heterogeneity, for example by including only individuals with amnestic, but not non-amnestic MCI (Jessen et al., 2018), we cannot exclude a potential influence of, for example, different atrophy patterns within the clinical groups (Baumeister et al., 2024). Ultimately, replication studies are needed to corroborate our findings, along with meta-analytic approaches with more stringent selection of contributing studies and contrasts (see Section 4.3).

Another more general limitation inherent to all fMRI studies in populations with cerebrovascular risk (e.g., aged populations, populations with AD/AD risk) is that changes of the cerebrovascular system can potentially affect the BOLD response (Sweeney et al., 2018; Zimmermann et al., 2021). Cerebral blood flow shortfalls are early findings in neurodegenerative disorders. Baseline differences in cerebral blood flow rates between experimental groups have the potential to produce a confound in the BOLD signal. However, in our study, we addressed specific contrasts rather than BOLD signal relative to baseline. Furthermore, differences between diagnostic groups do not only include reduced deactivations, but also atypial activations on the memory contrast (i.e., effects of subsequent memory) in DMN regions for Alzheimer’s disease patients (see Figure 1E in Soch et al., 2024). Therefore, we conclude that, although the potential impact of cerebrovascular differences cannot be excluded, it is, in our view, unlikely that vascular effects are the main drivers of our results. Furthermore, potential vascular contributions to the reduced expression of fMRI subsequent memory effects in individuals with MCI and AD do not call into question that the effects are reduced. Potential differences in cerebrovascular health therefore warrant caution with respect to mechanistic interpretations of our findings, but are unlikely to affect their potential diagnostic utility (see also Soch et al., 2024).

A limitation more specific to the present study is that participant groups significantly differed regarding age range, gender distribution, acquisition site (see Table 1), ApoE genotype and cognitive measures (MMSE total, NPT global and PACC5 scores; see Soch et al., 2024). While some of these differences are a direct consequence of the study design (e.g., AD patients show lower cognitive performance than the HC or SCD groups), other variables constitute confounds which cannot be as easily integrated into cvBMS as, for example, in a statistical design like a linear regression analysis.

We suggest that, to overcome at least some of the aforementioned limitations, future studies should assess the potential of reductionist or whole-brain multivariate data analysis approaches to both test for pathology-related deviations from more prototypical fMRI activations and assess the influence of potential risk factors (e.g., amyloid pathology) or confounding variables (e.g., study site). To this end, we have employed contrast maps obtained with the winning theoretical parametric GLM (i.e., the model using the arcsine-transformed memory regressor) to calculate single-value scores (Soch et al., 2021b; Richter et al., 2023). In a direct follow-up to the present study, we describe the extent to which these scores can further differentiate between the diagnostic groups in the clinical sample described here (Soch et al., 2024). Furthermore, we are currently working on improved computational modeling of the subsequent memory reports, which could be used in the future to differentiate participant groups based on purely behavioral response patterns (Soch et al., 2022).

### 4.5. Conclusions

Taken together, we could replicate the preference for parametric over categorical models of the fMRI subsequent memory effect in healthy older adults (Soch et al., 2021a) and demonstrate that this pattern also applies to cognitively unimpaired individuals at increased risk for Alzheimer’s disease (SCD, AD-rel). In individuals with MCI or manifest AD, on the other hand, memory-invariant models outperform any model considering the subsequent memory effect. Our results suggest that voxel-wise memory-related fMRI activity patterns in MCI or AD should be interpreted with caution and point to the need for additional or alternative analyses strategies, such as whole-brain approaches, in these populations.

## Data Availability

Data from the DELCODE study are available via individual data sharing agreements with the DELCODE study board (for more information, see https://www.dzne.de/en/research/studies/clinical-studies/delcode/). The code used for the Bayesian model selection of first-level fMRI data from the FADE paradigm has been published previously (Soch et al., 2021) and is available via GitHub (https://github.com/JoramSoch/FADE_BMS).

## 5. Notes

### 5.1. Ethics Statement

All participants and informants gave written informed consent to participate in the study in accordance with the Declaration of Helsinki. The DELCODE study protocol was approved by the ethics committees of the medical faculties of all recruiting sites: Berlin (Charité, University Medicine), Bonn, Cologne, Göttingen, Magdeburg, Munich (Ludwig-Maximilians-University), Rostock, and Tübingen. The ethics approval process was coordinated by the ethics committee of the medical faculty of the University of Bonn (registration number 117/13). DELCODE was registered as a clinical trial with the German Clinical Trials Register (https://www.bfarm.de/EN/BfArM/Tasks/German-Clinical-Trials-Register/_node.html) under the study ID DRKS00007966.

### 5.2. Data and code availability

Data from the DELCODE study are available via individual data sharing agreements with the DELCODE study board (for more information, see https://www.dzne.de/en/research/studies/clinical-studies/delcode/). The code used for the Bayesian model selection of first-level fMRI data from the FADE paradigm has been published previously (Soch et al., 2021b) and is available via GitHub (https://github.com/JoramSoch/FADE_BMS).

## 5.3. Acknowledgments

We would like to thank all the participants in the DELCODE study and of the *Autonomy in Old Age* study and all the technical, medical and psychological staff for making this study possible. Special thanks go to the Max Delbrück Center for Molecular Medicine (MDC) within the Helmholtz Association, the Center for Cognitive Neuroscience Berlin (CCNB) at the Free University of Berlin, the Bernstein Center for Computational Neuroscience (BCCN) Berlin, the MR research core facility of the University Medical Center Göttingen (UMG) and the MR research center of the University Hospital Tübingen (UKT).

## 6. Appendix

**Figure 8.**
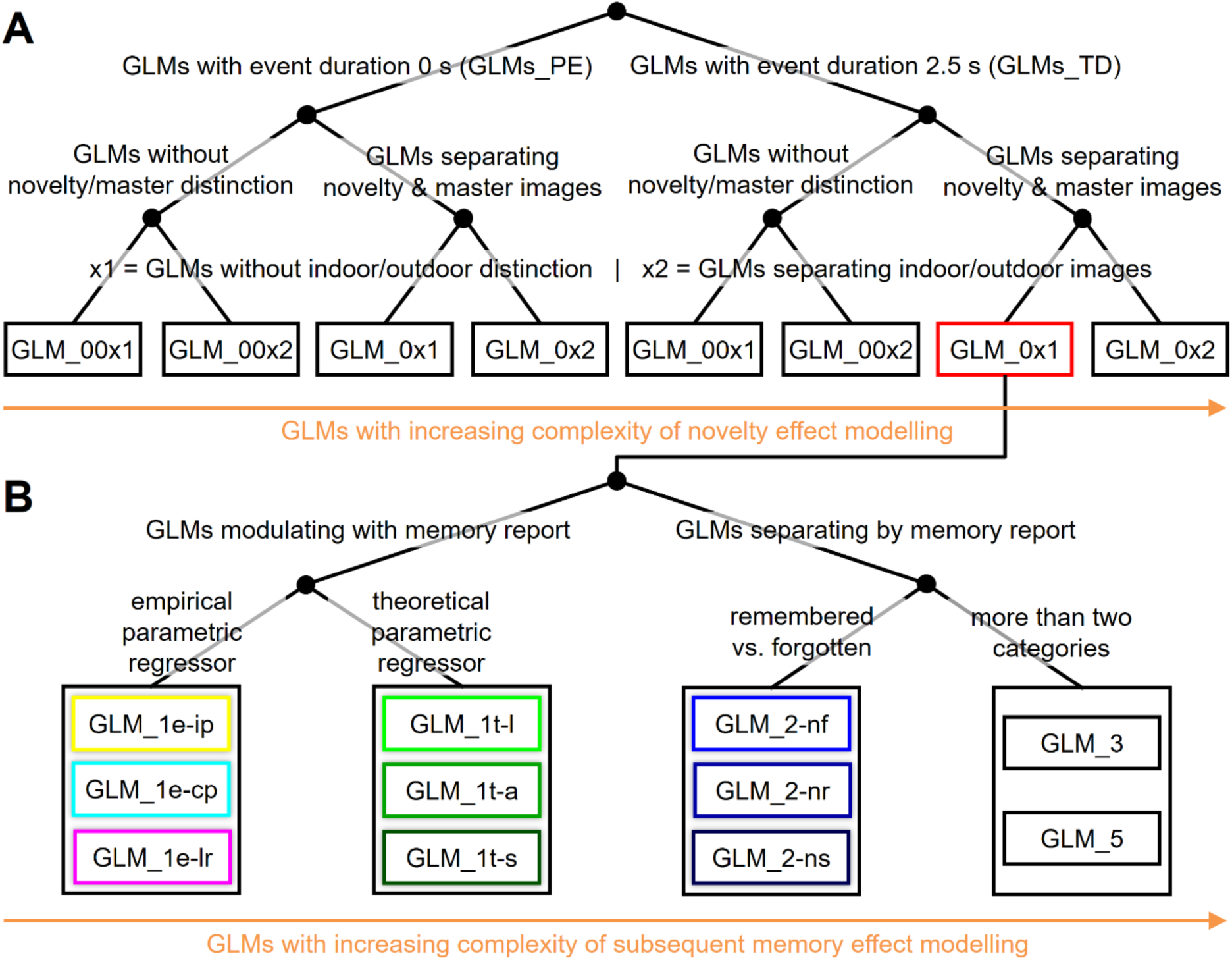
Model space for GLM-based fMRI analyses. **(A)** 8 models without memory effects varying model features of no interest, namely modeled event duration (top), consideration of stimulus novelty (middle) and consideration of stimulus type (bottom). **(B)** 11 models varying by the way how memory effects are modeled. Each box represents a single first-level GLM; the box with red outline represents the model referred to as “baseline GLM” in this paper. This figure reproduces Supplementary Figure S2 from the original publication which is available under the license CC-BY-NC-ND 4.0 (original work at: https://www.sciencedirect.com/science/article/pii/S1053811921000975#sec0031; license file at: https://creativecommons.org/licenses/by-nc-nd/4.0/).

**Table 3.**
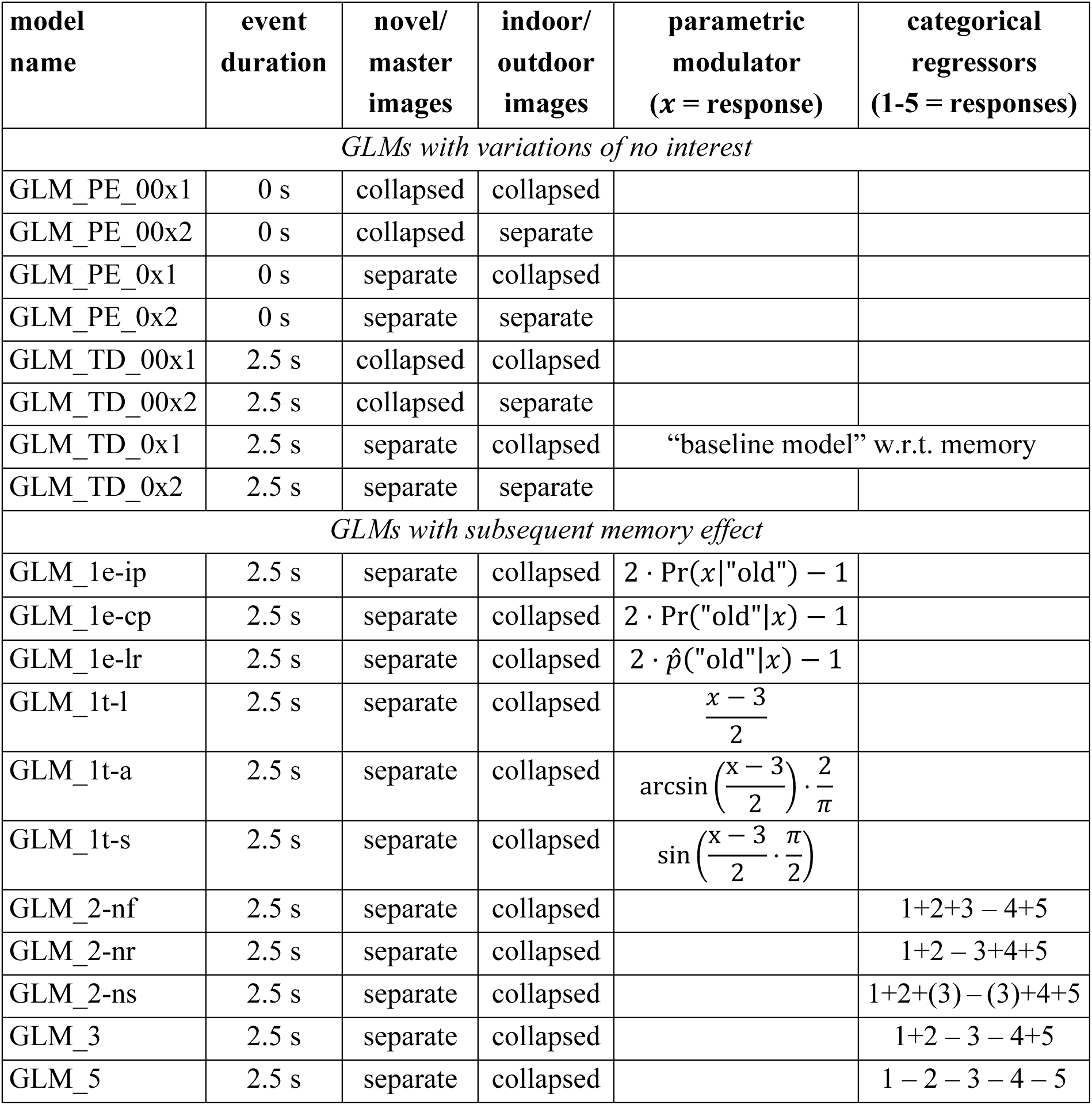
Model space for GLM-based fMRI analyses. 8 models without memory effects varying model features of no interest (top) and 11 models varying by the way how memory effects are modelled (bottom). All parametric modulators are specified, such that the categorical responses {1, 2, 3, 4, 5} are mapped into the range [–1, +1]. This table reproduces Table 1 from the original publication which is available under the license CC-BY-NC-ND 4.0 (original work at: https://www.sciencedirect.com/science/article/pii/S1053811921000975#tbl0001; license file at: https://creativecommons.org/licenses/by-nc-nd/4.0/).

## Footnotes

1 https://www.dzne.de/en/research/studies/clinical-studies/delcode/

2 The DELCODE proposal for this data analysis (DELCODE 243) is available from the authors upon request.

3 https://www.fil.ion.ucl.ac.uk/spm/software/spm12/

4 Note that a thorough statistical analysis of the between-group differences in behavioural response frequencies and subsequent memory reports will be the focus of a later publication (see Soch et al., 2022 for methodology).

5 Note that novelty contrasts from other models give rise to very similar results, since memory models of interest did not differ in their description of the novelty effect.

