## supplementary text, figures, and tables for "Reduced expression of fMRI subsequent memory effects with increasing severity across the Alzheimer’s disease risk spectrum"

### Supplementary Methods

##### Extended figures for Bayesian model comparison and statistical significance testing

The presentation of results in this supplementary material is as follows: For each analysis conducted in the original study (Soch et al., 2021; see Table S1), we present an extended figure with mostly five sagittal slices as an in-depth view of model preferences and statistical significance (e.g. Figures S7-S9; corresponding to single-slice results on Figure 5 in the main manuscript). This includes analyses whose results are already shown in reduced form in the main manuscript (e.g. Figure S4; corresponding to Figure 3A in the original study; extending results from Figure 2B in the main manuscript) as well as analyses whose results are only shown in this supplementary material (e.g. Figure S1; corresponding to Figure S3A in the original study; not shown in the main manuscript). An index of all analyses can be found in Table S1.

##### Cluster-level correction of group-level statistical analyses

For analysis of novelty processing and subsequent memory effects in each participant group (see Figure 7), we additionally perform cluster-level correction instead of the whole-brain family-wise error (FWE) correction used in the main manuscript, as requested by Reviewer 2. We use a cluster-defining threshold (CDT) of $p < 0.001$ which resulted in different cluster extent thresholds ($FWE_c$) for novelty models (HC: $k = 67$; SCD: $k = 80$; MCI: $k = 33$; AD: $k = 146$; AD-rel: $k = 36$) and memory models (HC: $k = 61$; SCD: $k = 34$; MCI: $k = 35$; AD: $k = Inf$; AD-rel: $k = 46$) that were applied for thresholding uncorrected maps (see Figure S14).

### Supplementary Results

#### Extended results from Bayesian model selection and statistical significance testing

Results not reported in the main manuscript, but only contained in this supplementary material, show that

- modelling encoding trials with an event duration of duration 2.5 s outperforms modelling them as point events of duration 0 s (see Figure S1);
- modelling indoor and outdoor images separately outperforms not doing so only in parts of occipital cortex not related to episodic memory (see Figure S3);
- including a novelty effect into the GLM also improves model quality in the bilateral temporo-parietal junction (TPJ; see Figure S2, outer-most slices);
- including a memory effect into the GLM also improves modal quality in the right TPJ in healthy controls, SCD patients and AD relatives (see Figure S4, right-most slices);
- model preferences within model families are not restricted to the center slice shown in the main manuscript (see Figure 5), but are uniform across the brain (see Figures S7-S9);
- group-level effects of subsequent memory obtained with the winning empirical parametric GLM (see Figure S12; inverse probability regressor) and using the winning two-regressor GLM (see Figure S13; remembered vs. forgotten) are qualitatively similar to those obtained using the winning theoretical parametric (and by us, recommended) GLM (see Figure 6B; arcsine-transformed regressor), albeit a bit weaker.

#### Group-level novelty and memory effects in MCI and AD patients

When comparing cluster-level-corrected to whole-brain FWE-corrected results for significant novelty and memory effects (see Figure 7 in the main manuscript), we additionally observe mild effects of novelty processing in the bilateral temporo-occipito-parietal network for AD patients (see Figure S14A, 4th row) and mild effects of subsequent memory in the precuneus und posterior cingulum for MCI patients (see Figure S14B, 3rd row). There is still no effect of subsequent memory for individuals with AD when correcting for cluster extent.

#### Supplementary Tables

| Analysis | Comparison | original study | present paper | this supplement |
|---|---|---|---|---|
| *Bayesian model selection* | | | | |
| 1 | GLMs with trial duration of 2.5 s vs. GLMs with trials modelled as point events | Figure S3A | – | Figure S1 |
| 2 | GLMs separating novel/master images vs. GLMs collapsing these events | Figure S3B | Figure 2/3A Table 2 | Figure S2 Table S3 |
| 3 | GLMs separating indoor/outdoor scenes vs. GLMs collapsing these stimuli | Figure S3C | – | Figure S3 |
| 4a | GLMs with memory effect vs. baseline GLM | Figure 3A | Figure 2/3B Table 2 | Figure S4 Table S3 |
| 4b | GLMs with 1 vs. 2 vs. 3 vs. 5 regressors | Figure 3B | Figure 4 | – |
| 5 | parametric vs. categorical memory GLMs | Figure 4A | Figure 5A | Figure S5 |
| 6 | empirical vs. theoretical parametric GLMs | Figure 4B | Figure 5B | Figure S6 |
| 7 | winning GLM with two memory regressors | Figure 5A | Figure 6A | Figure S7 |
| 8 | winning theoretical parametric GLM | Figure 5B | Figure 6B | Figure S8 |
| 9 | winning empirical parametric GLM | Figure 5C | Figure 6C | Figure S9 |
| 10a | winning empirical parametric GLM vs. winning theoretical parametric GLM | Figure S4A | – | – |
| 10b | winning theoretical parametric GLM vs. winning empirical parametric GLM | Figure S4B | – | – |
| *Statistical significant testing* | | | | |
| A | novelty contrast from GLM_1t-a | Figure 7A | Figure 7A | Figure S10 |
| B | memory regressor from GLM_1t-a | Figure 7B | Figure 7B | Figure S11 |
| C | memory regressor from GLM_1e-ip | Figure 7C | – | Figure S12 |
| D | memory contrast from GLM_2nf | Figure 7D | – | Figure S13 |

**Table S1.** *Index of group-level fMRI analyses.* This table lists group fMRI analyses conducted in the original study and replicated for the present paper. The analysis IDs in the first column refers to the IDs used in the original study. Models described in the second column refer to those describe in Table 3 in the Appendix of the main manuscript. The last three columns list where to find results in the original paper, in the main manuscript and in this supplement.

| Step of data acquisition/processing | Description in Soch et al., 2021 |
|---|---|
| experimental paradigm | see Section 2.2 and Figure 1 |
| fMRI data acquisition | see Section 2.3 |
| fMRI data preprocessing | see Section 2.4 |
| general linear modelling | see Section 3 and Figure 2 |
| model space of GLMs | see Section 3 and Table 1 |
| Bayesian model selection | see Section 2.5 |

**Table S2.** *Reference for data acquisition and processing.* Steps of data acquisition and processing are summarized in Sections 2.3 to 2.6 of the main manuscript. Details can be found in the referenced sections of the original publication (right column).

| | novelty processing | subsequent memory |
|---|---|---|
| main effect of diagnosis | $F_{4,446} = 4.00$, $p = 0.003$ | $F_{4,442} = 3.64$, $p = 0.006$ |
| main effect of gender | $F_{1,446} = 14.99$, $p < 0.001$ | $F_{1,442} = 1.86$, $p = 0.174$ |
| interaction of diagnosis and gender | $F_{4,446} = 1.85$, $p = 0.118$ | $F_{4,442} = 0.37$, $p = 0.827$ |
| main effect of site | $F_{7,446} = 2.27$, $p = 0.028$ | $F_{7,442} = 2.39$, $p = 0.021$ |
| effect of age | $F_{1,446} = 19.53$, $p < 0.001$ | $F_{1,442} = 1.61$, $p = 0.205$ |
| effect of educational years | $F_{1,446} = 0.66$, $p = 0.417$ | $F_{1,442} = 0.15$, $p = 0.698$ |
| effect of employment years | $F_{1,446} = 0.07$, $p = 0.787$ | $F_{1,442} = 0.10$, $p = 0.755$ |

**Table S3.** *Analysis of covariance for number of voxels selected via Bayesian model selection.* Number of voxels exceeding a log Bayes factor of 3 (approximately, a Bayes factor of 20) in Bayesian model comparisons testing for novelty processing and subsequent memory (cf. Figure 2 and Table 2) were extracted for all participants from all diagnostic groups. Each cell reports F-statistic and p-value for categorical effects of factors (diagnosis, gender, site) or parametric effects of covariates (age, education, employment) on number of voxels, estimated from an analysis of covariance (ANCOVA) model jointly including all these variables. Indices of F-statistics indicate numerator and denominator degrees of freedom. This Table lists test statistics for variables that were controlled for when reporting main effects of diagnosis in Figure 2.

#### Supplementary Figures

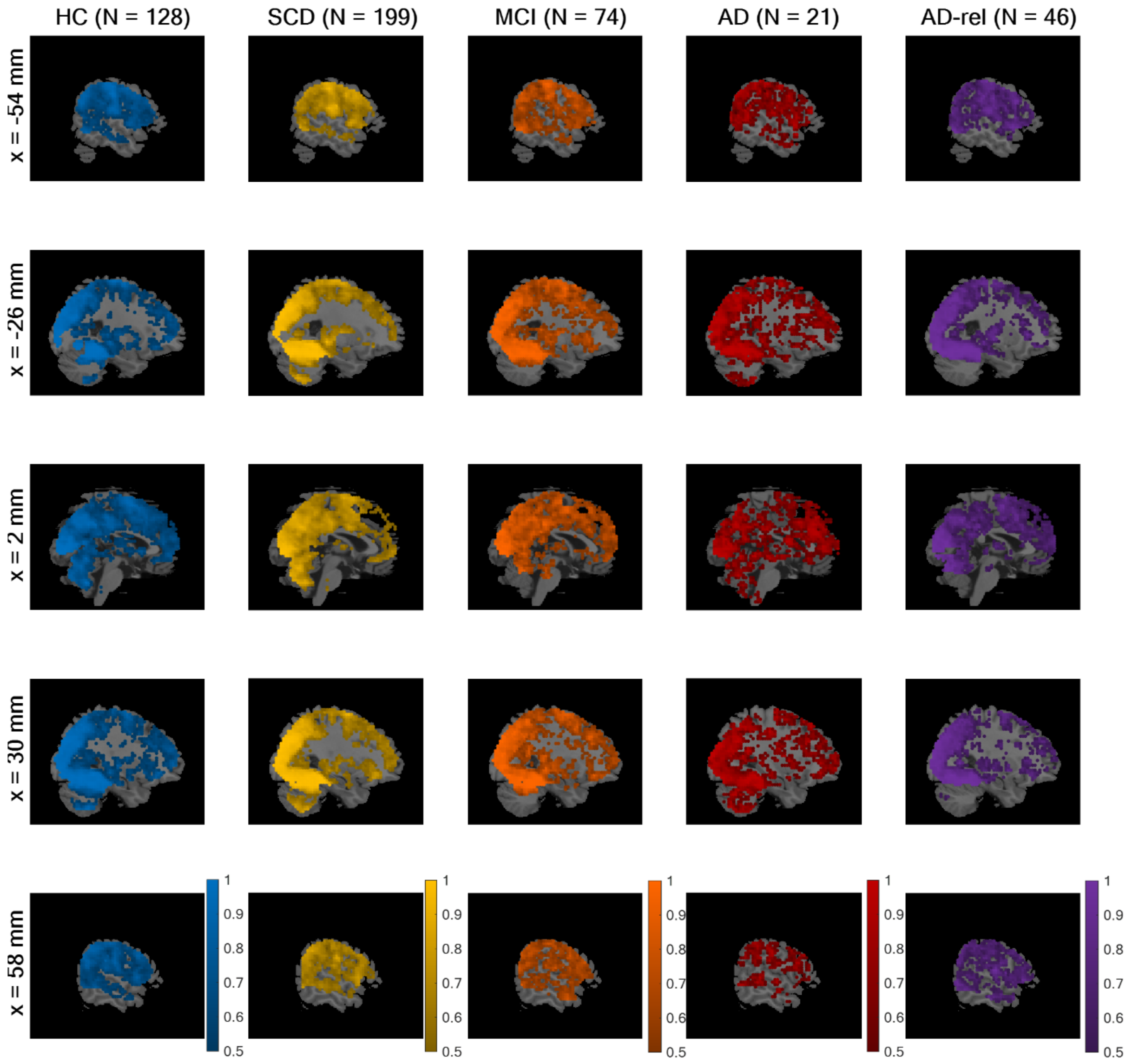


**Figure S1.** *Effects of event duration.* Selected-model maps, showing voxels with group-level preference in favor of the family of models using the trial duration, against the family of models assuming point events. Five sagittal slices (x-coordinate given on top), roughly equal to those used in results display in the original study, are shown for each subject group (sample size given at the left) and colored voxels indicate a higher estimated frequency of the model family "GLMs_TD", rather than the model family "GLMs_PE". This figure corresponds to Figure S3A from the original publication.

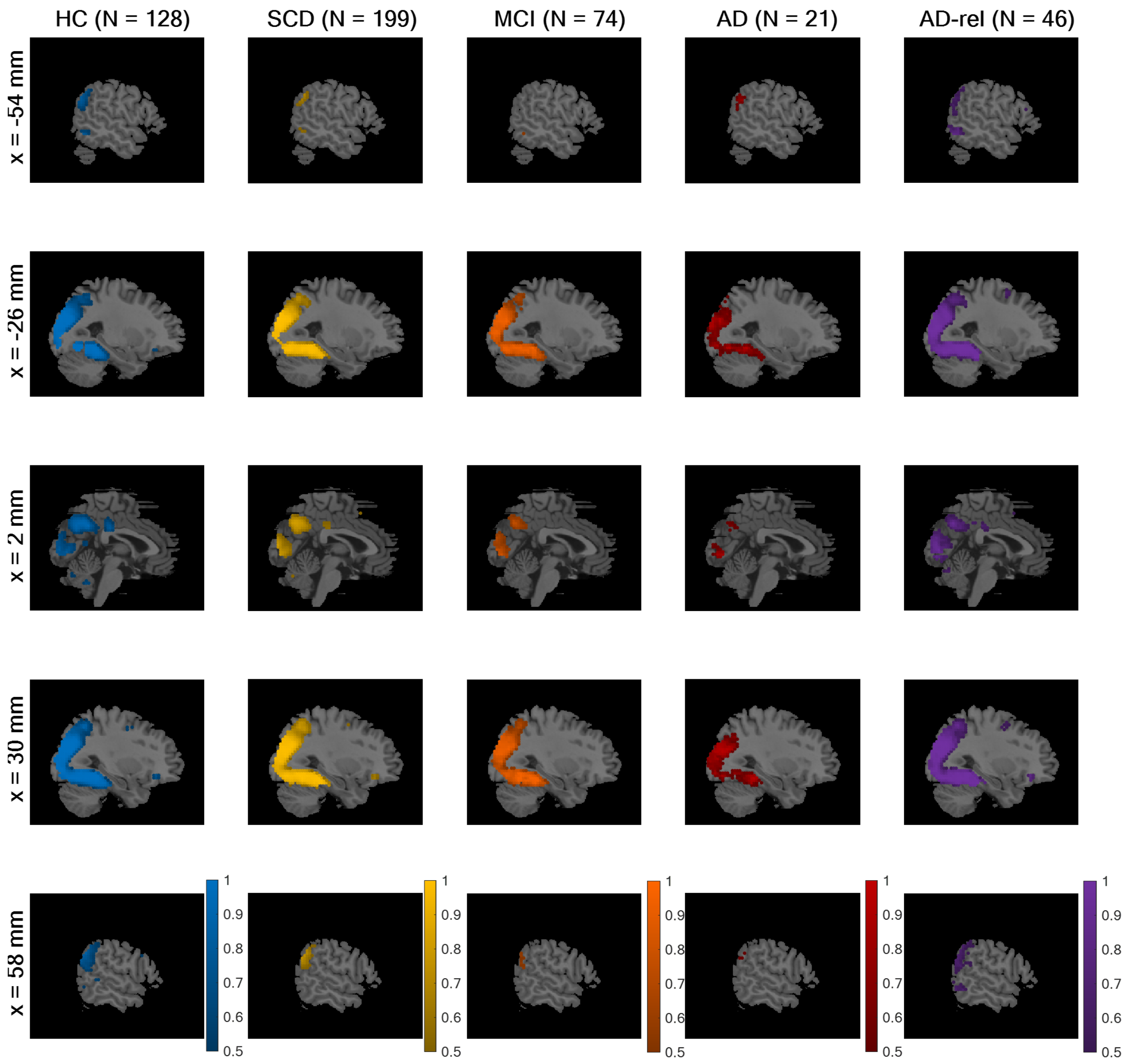


**Figure S2.** *Effects of novelty processing.* Selected-model maps, showing voxels with group-level preference in favor of the family of models separating novel and pre-familiarized images, against the family of models not considering novelty. The layout of the figure follows that of Figure S1. Colored voxels indicate a higher estimated frequency of the model family "GLMs_0" (novelty and master regressor), rather than the model family "GLMs_00" (both regressors collapsed). This figure corresponds to Figure S3B from the original publication.

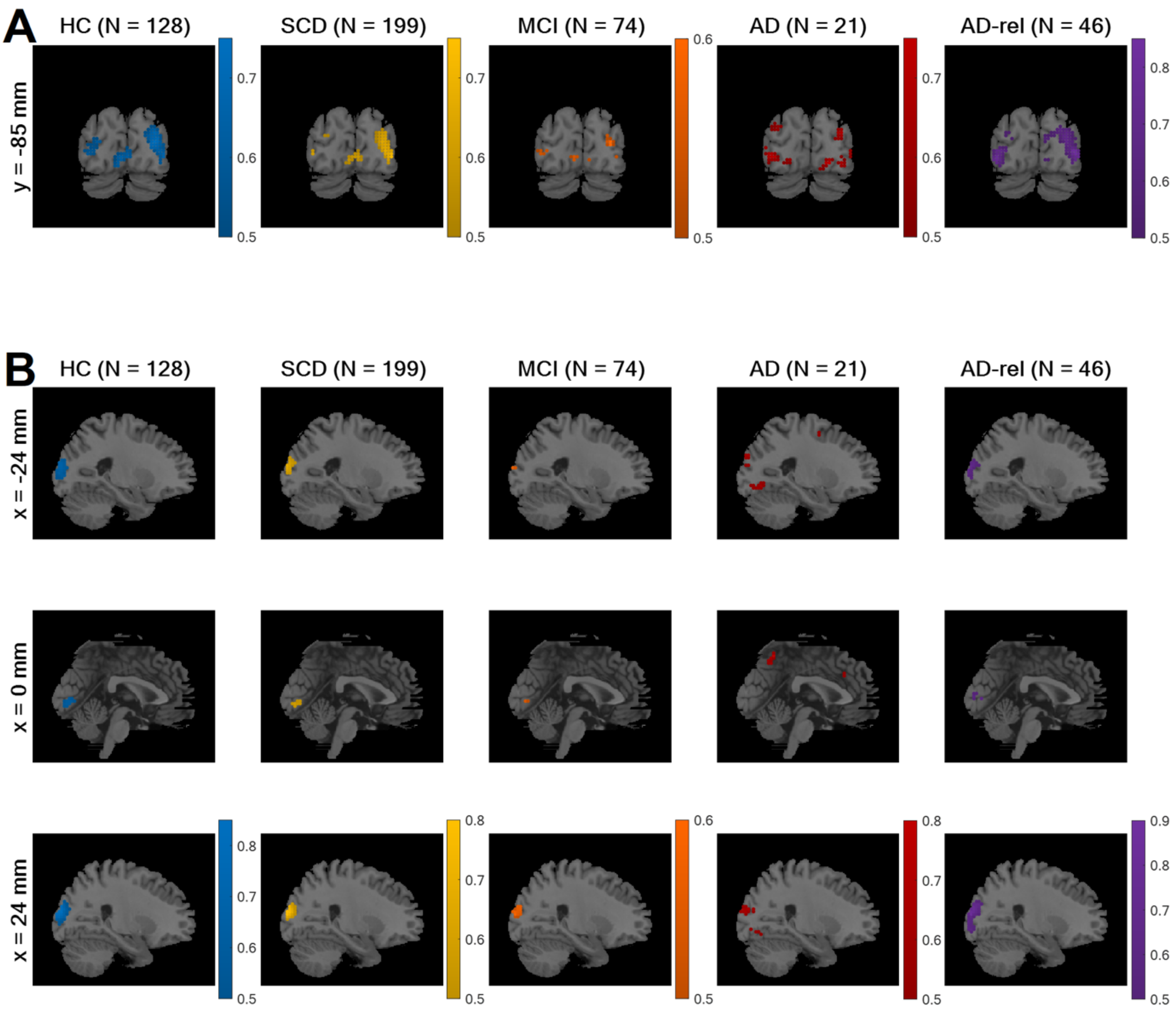


**Figure S3.** *Effects of stimulus type.* Selected-model maps, showing voxels with group-level preference in favor of the family of models separating indoor and outdoor scenes, against the family of models not doing so. **(A)** One coronal slice (y-coordinate given on top) and **(B)** three sagittal slices (x-coordinates given on top), roughly equal to those used in results display in the original study, are shown for each subject group (sample size given at the left). Colored voxels indicate a higher estimated frequency of the model family "GLMs_x2" (indoor and outdoor regressors), rather than the model family "GLMs_x1" (both categories collapsed). This figure corresponds to Figure S3C from the original publication.

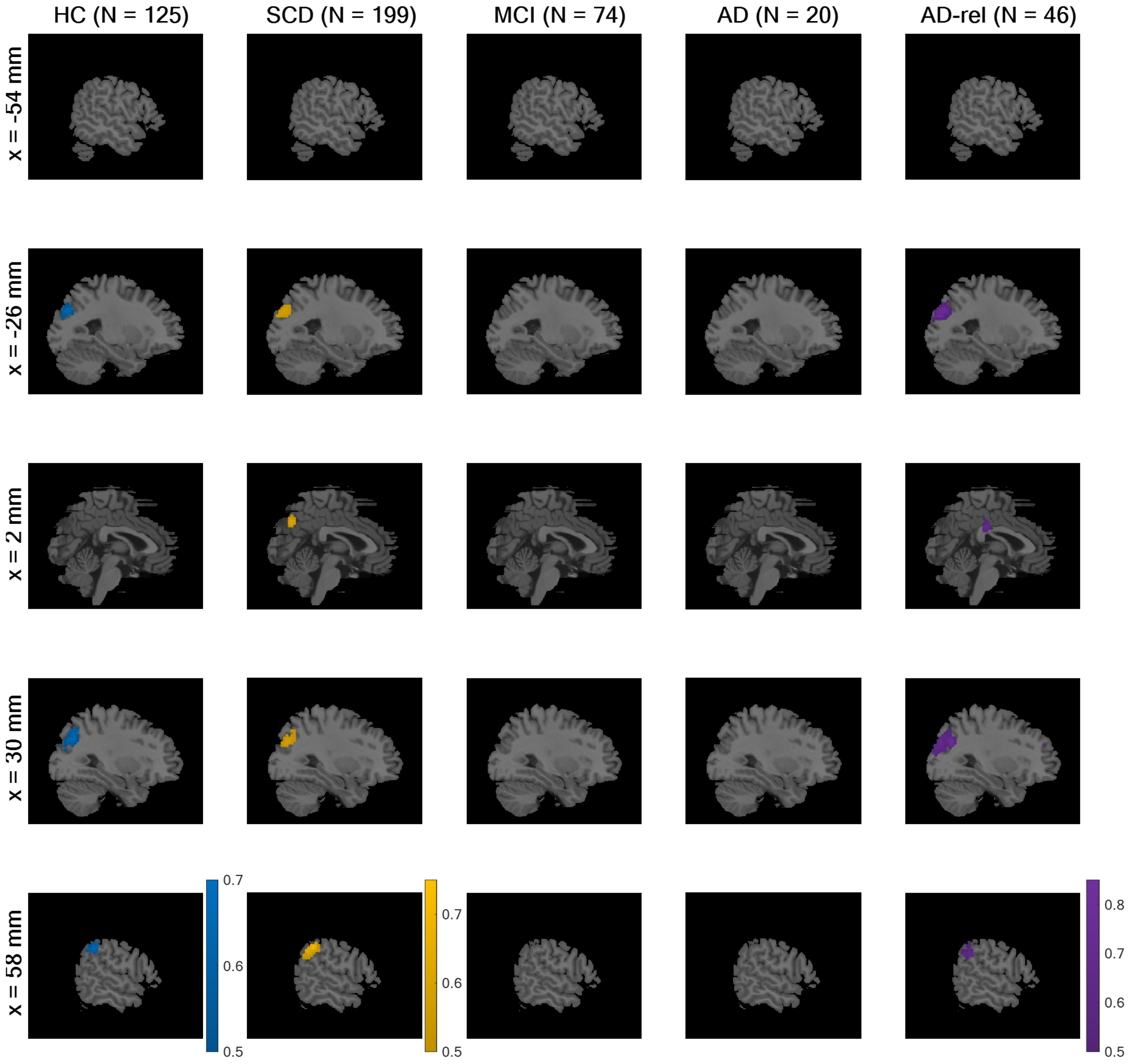


**Figure S4.** *Effects of subsequent memory.* Selected-model maps, showing voxels with group-level preference in favor of memory models, against the baseline GLM. The layout of the figure follows that of Figure S1. Colored voxels indicate a higher estimated frequency of the model family "GLMs_12" (one or two memory regressors), rather than the model "GLM_TD_0x1" (no memory effect). This figure corresponds to Figure 3A from the original publication.

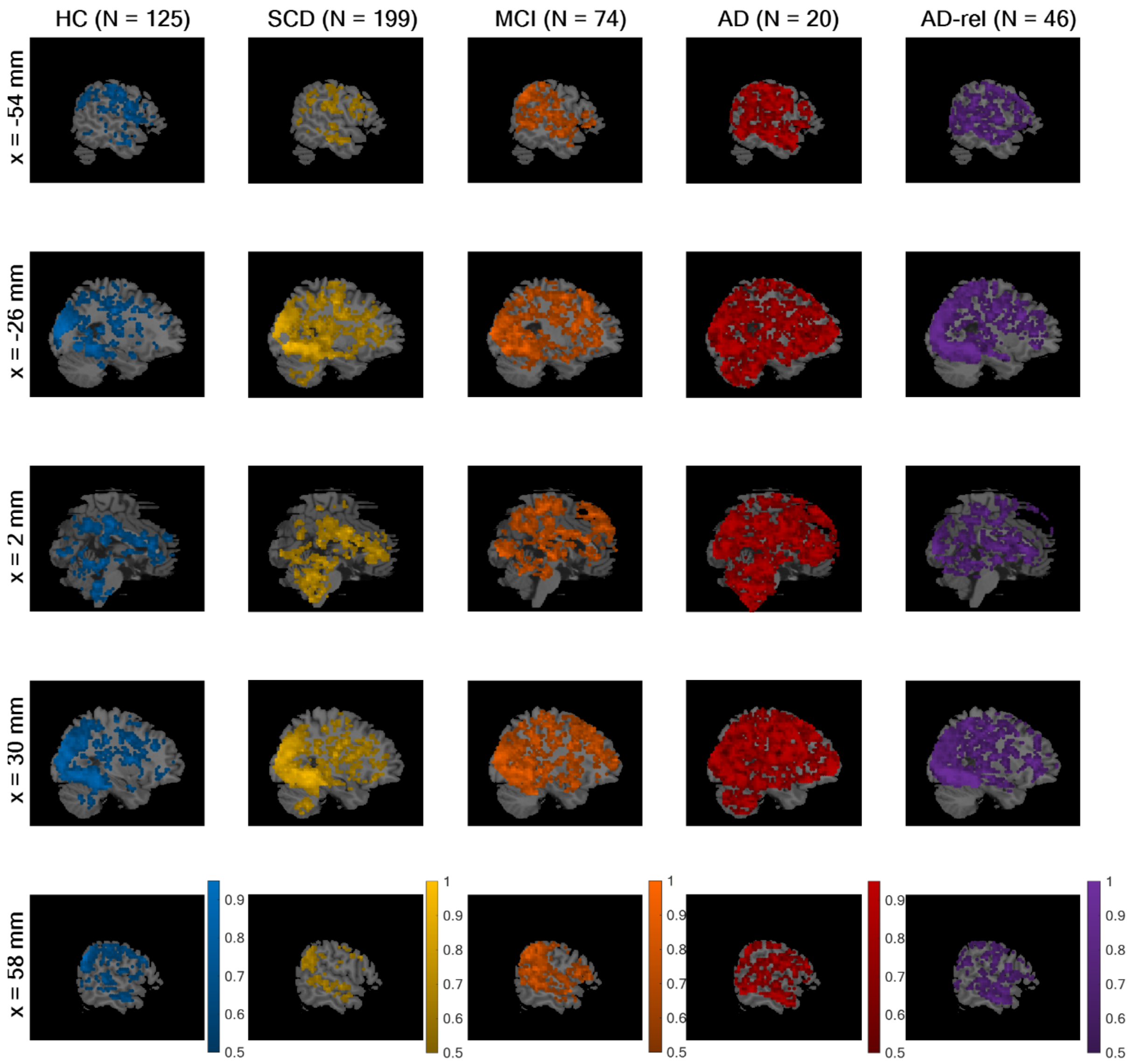


**Figure S5.** *Parametric vs. categorical memory models.* Selected-model maps, showing voxels with group-level preference in favor of parametric models of the subsequent memory effect, using parametric modulators, against categorical models, separating response options. The layout of the figure follows that of Figure S1. Colored voxels indicate a higher estimated frequency of the model family "GLMs_1" (one memory regressor), rather than the model family "GLMs_2" (two memory regressors). This figure corresponds to Figure 4A from the original publication.

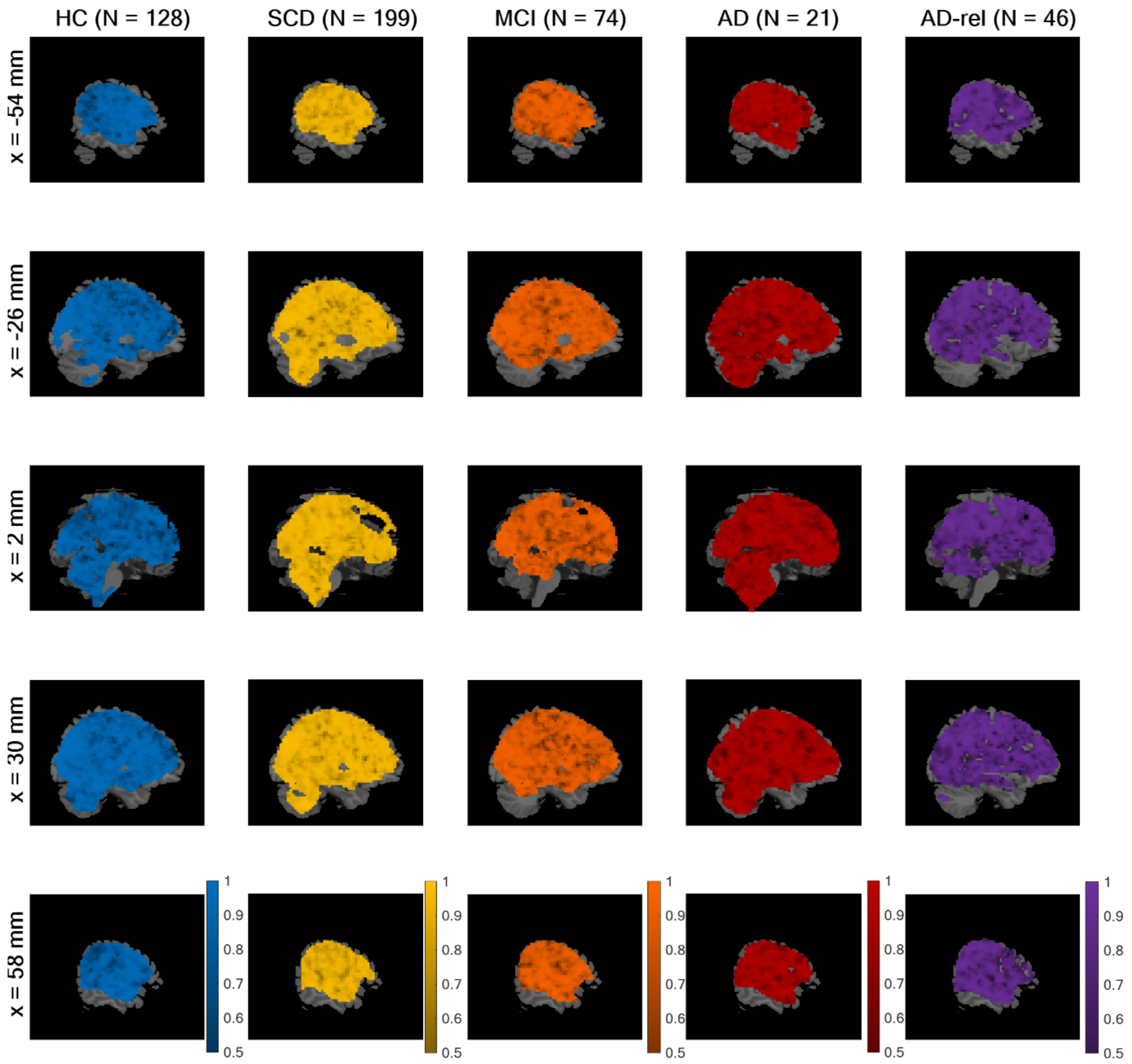


**Figure S6.** *Empirical vs. theoretical parametric models.* Selected-model maps, showing voxels with group-level preference in favor of empirical parametric models, using data-driven transformations, against theoretical models, using *a priori* defined transformations. The layout of the figure follows that of Figure S1. Colored voxels indicate a higher estimated frequency of the model family "GLMs_1e" (empirical modulators), rather than the model family "GLMs_1t" (theoretical modulators). This figure corresponds to Figure 4B from the original publication.

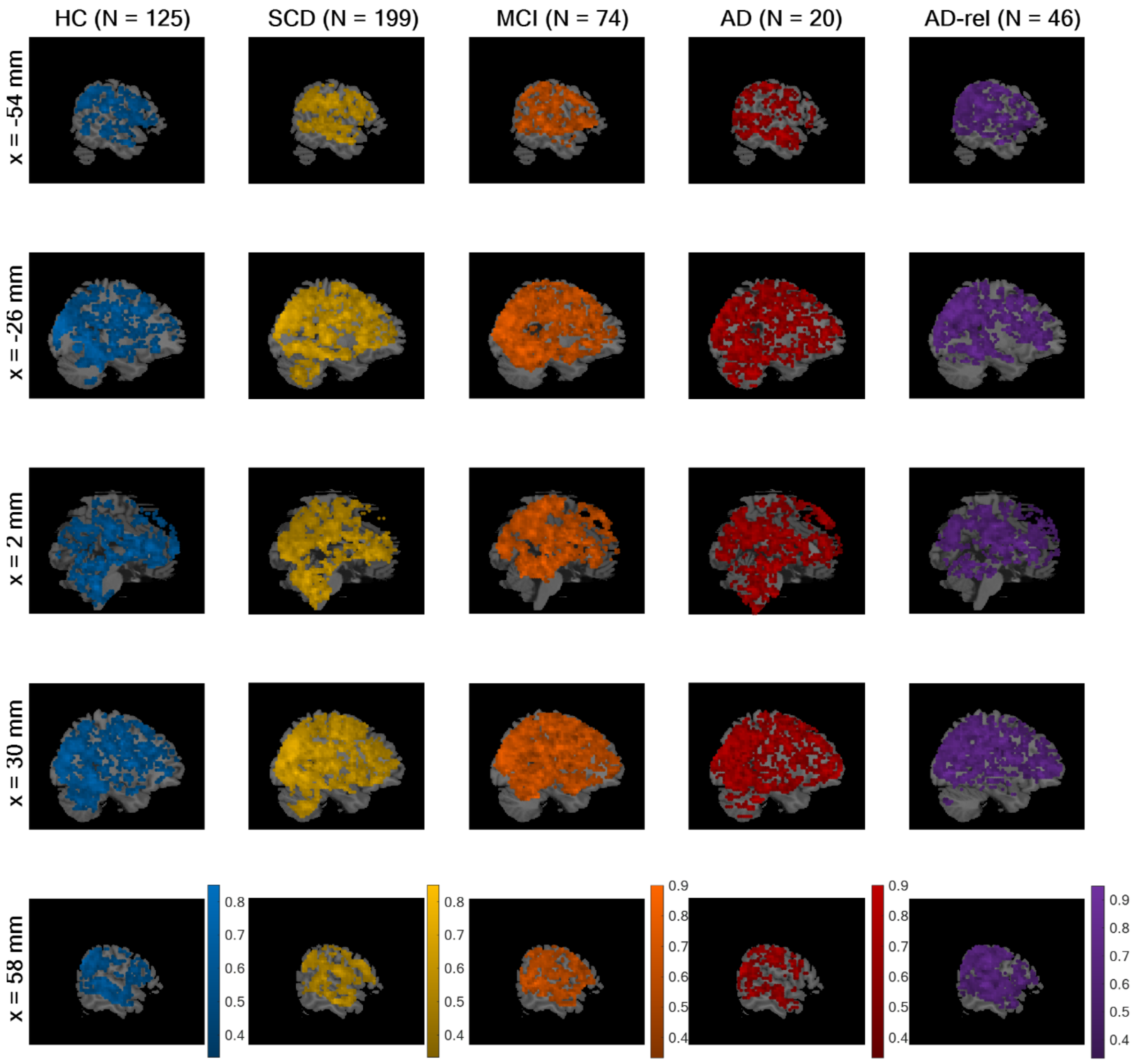


**Figure S7.** *Winning models within two-regressor family.* Selected-model maps in favor of the GLM treating neutral images as forgotten items ("GLM_2-nf"), compared to the GLM treating neutral images as remembered items ("GLM_2-nr") and the GLM randomly assigning neutral images to remembered or forgotten items ("GLM_2-ns"). The layout of the figure follows that of Figure S1. This figure corresponds to Figure 5A from the original publication.

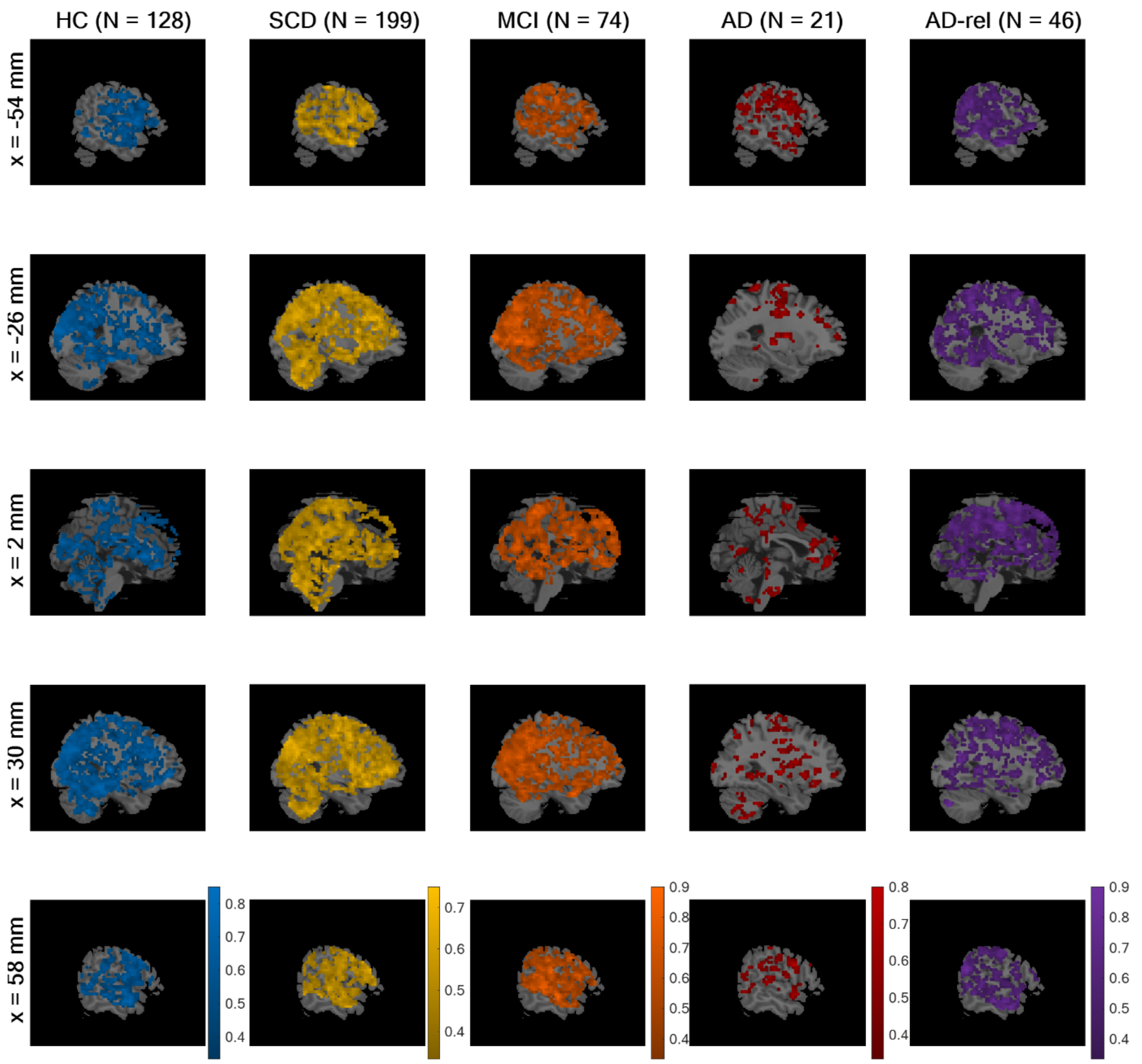


**Figure S8.** *Winning models within theoretical parametric GLMs.* Selected-model maps in favor of the GLM using an arcsine-transformed parametric modulator ("GLM_1t-a"), compared to the GLM using a sine-transformed parametric modulator ("GLM_1t-s") and the GLM using a linear parametric modulator ("GLM_1t-l"). The layout of the figure follows that of Figure S1. This figure corresponds to Figure 5B from the original publication.

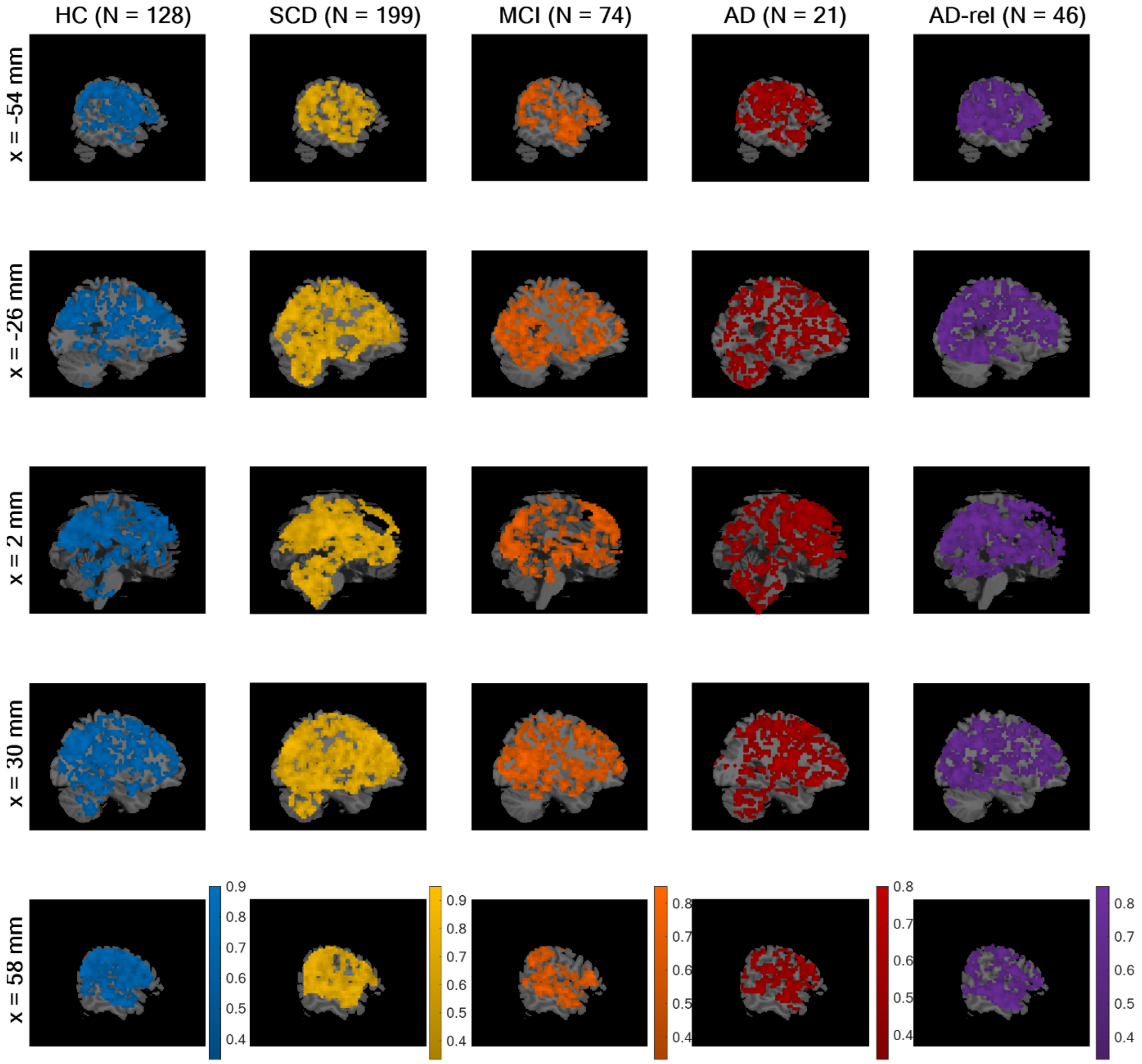


**Figure S9.** *Winning models within empirical parametric GLMs.* Selected-model maps in favor of the GLM using an inverse probability parametric modulator ("GLM_1e-ip"), compared to the GLM using a conditional probability parametric modulator ("GLM_1e-cp") and the GLM using a parametric modulator based on logistic regression ("GLM_1e-lr"). The layout of the figure follows that of Figure S1. This figure corresponds to Figure 5C from the original publication.

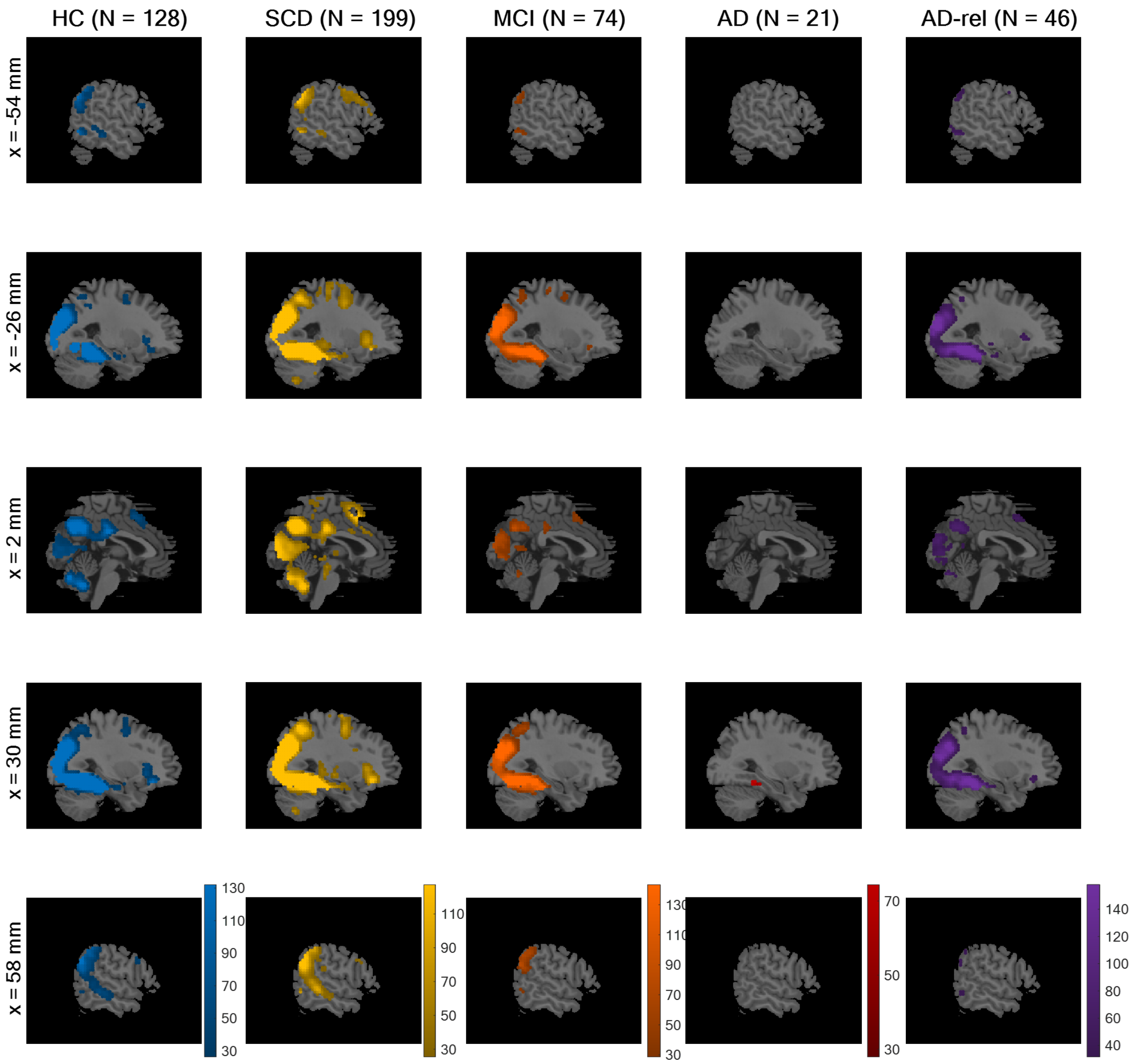


**Figure S10.** *Novelty contrast from winning theoretical parametric GLM.* On the second level, a one-sample t-test was run across parameter estimates obtained from the novelty contrast (novel vs. master images) of the GLM using the arcsine-transformed PM. In SPM, an F-contrast was specified on the single mean parameter and statistical inference was corrected for multiple comparisons (FWE, $p < 0.05$, $k = 10$). Colored voxels indicate significant differences between novel and master images, on average across subjects from the respective participant group. This figure corresponds to Figure 7A from the original publication.

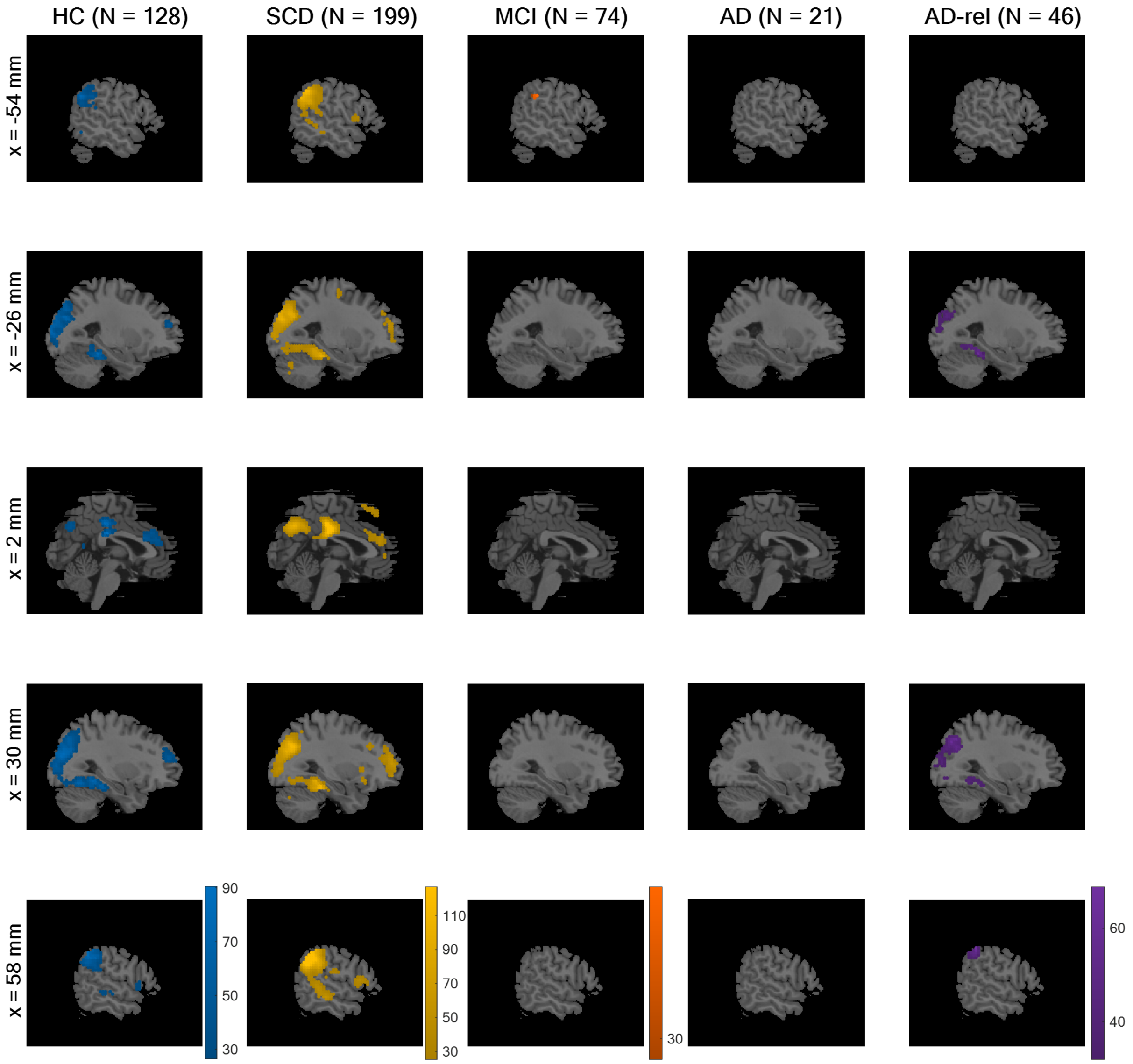


**Figure S11.** *Memory regressor from winning theoretical parametric GLM.* On the second level, a one-sample t-test was run across parameter estimates obtained from the memory regressor of the theoretical parametric GLM using the arcsine-transformed PM. The layout of the figure follows that of Figure S10. Colored voxels indicate significant non-zero effect of transformed memory response, on average across subjects from the respective participant group. This figure corresponds to Figure 7B from the original publication.

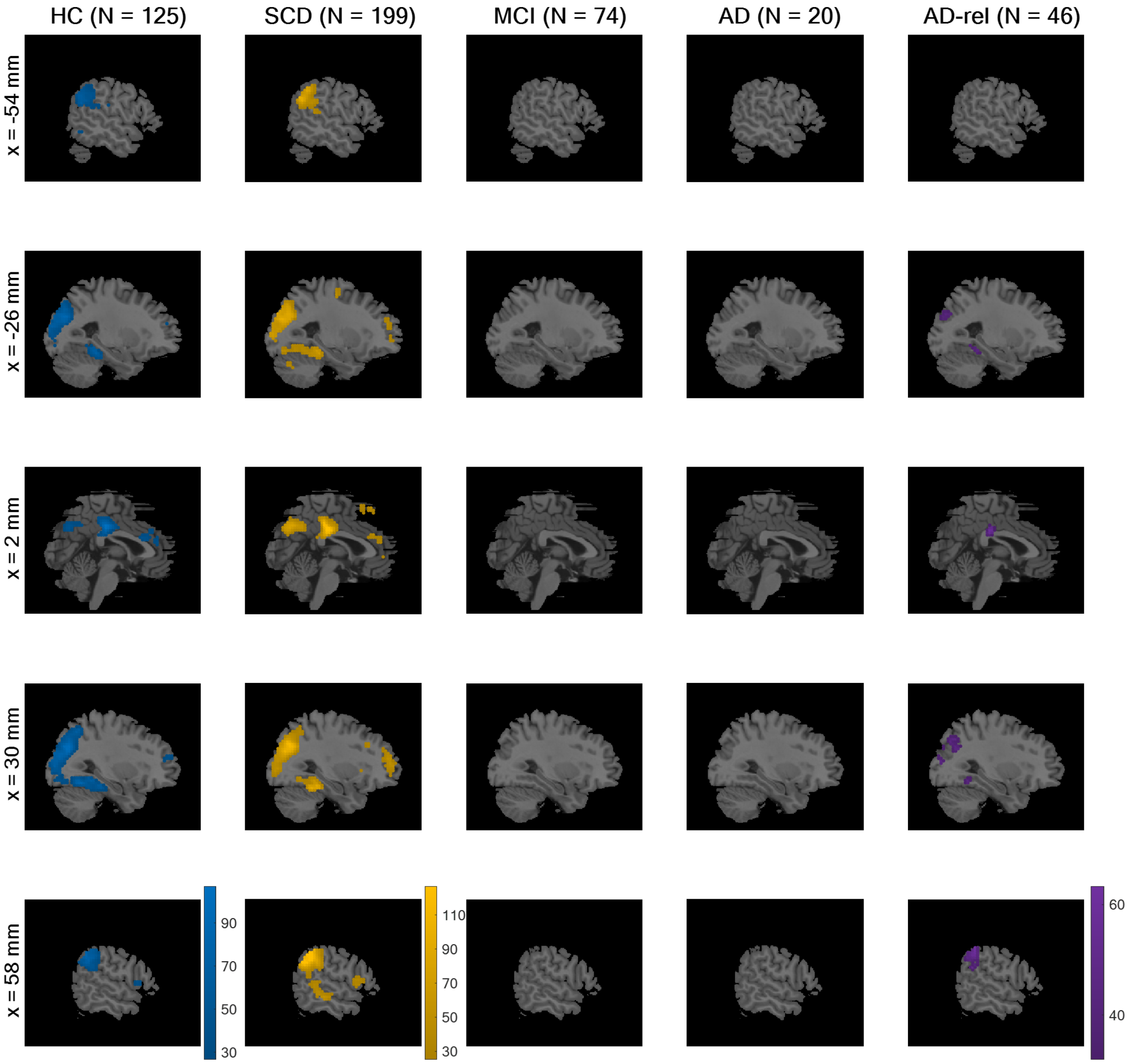


**Figure S12.** *Memory regressor from winning empirical parametric GLM.* On the second level, a one-sample t-test was run across parameter estimates obtained from the memory regressor of the empirical parametric GLM using the inverse probability PM. The layout of the figure follows that of Figure S10. Colored voxels indicate significant non-zero effect of normalized inverse probability, on average across subjects from the respective participant group. This figure corresponds to Figure 7C from the original publication.

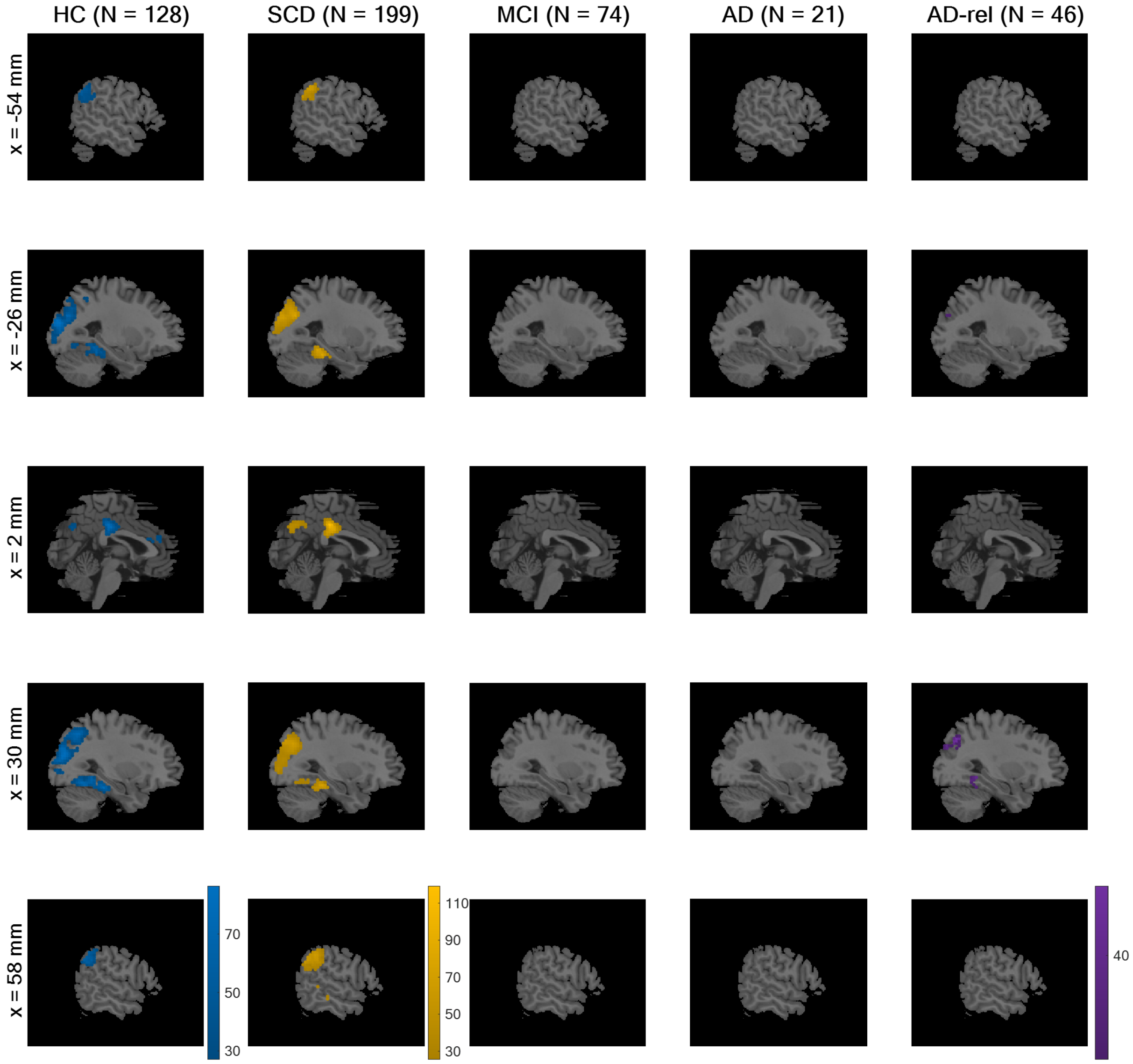


**Figure S13.** *Memory contrast from winning two-regressor model.* On the second level, a one-sample t-test was run across parameter estimates obtained from the memory contrast (remembered vs. forgotten items) resulting from the categorical GLM categorizing neutral responses as forgotten. The layout of the figure follows that of Figure S10. Colored voxels indicate significant differences between remembered and forgotten items, on average across subjects from the respective participant group. This figure corresponds to Figure 7D from the original publication.

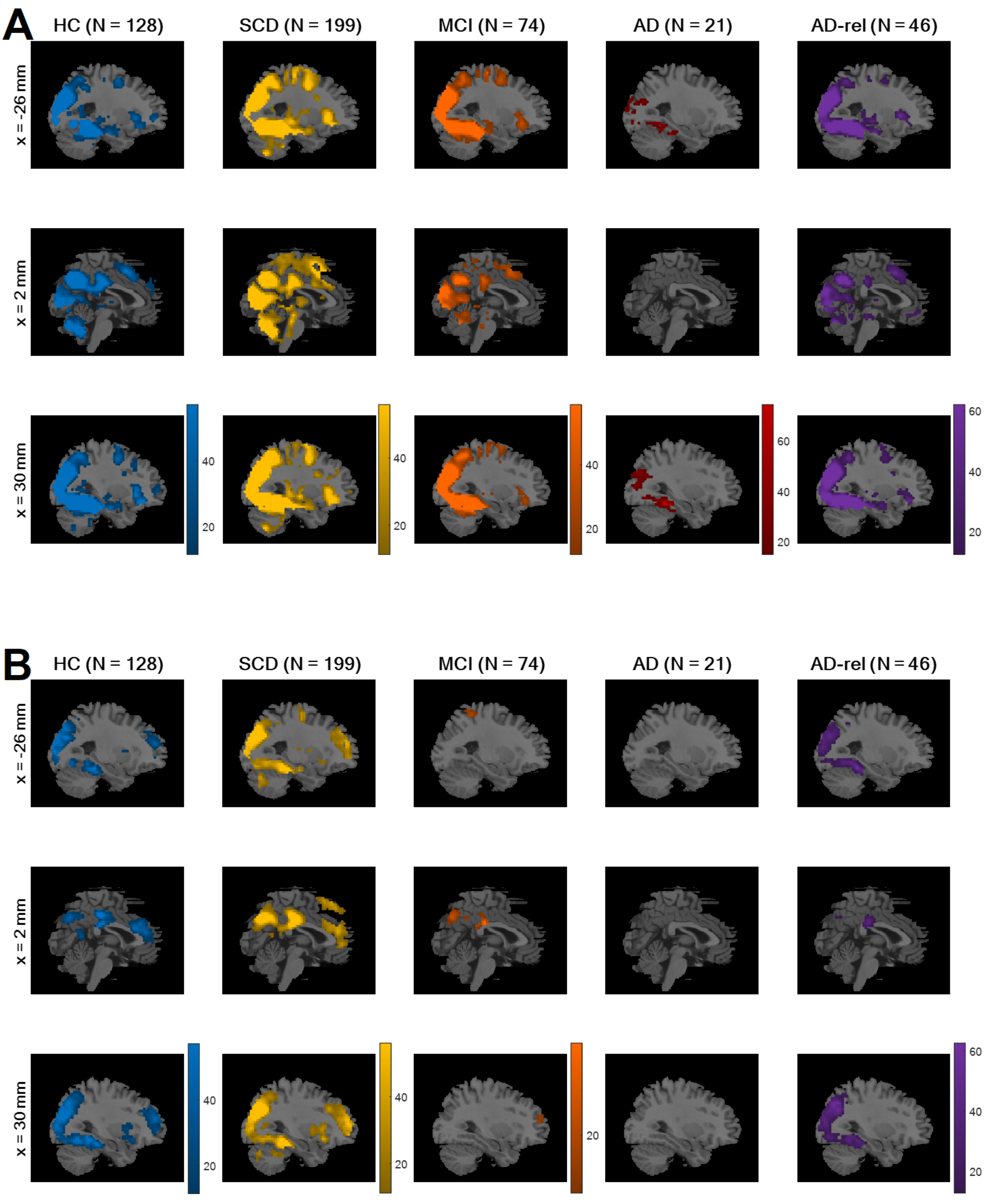


**Figure S14.** *Novelty and memory effects from winning GLM (cluster-corrected).* Colored voxels display F-statistics indicating **(A)** significant (positive or negative) differences between novel and master images and **(B)** significant (non-zero) effects of the transformed memory response, on average across subjects from the respective participant group. In SPM, statistical inference was cluster-level-corrected for family-wise error (FWE) using a cluster-defining threshold (CDT) of $p < 0.001$, resulting in different cluster extent thresholds (see Methods). This figure represents a cluster-corrected version of Figure 7 in the main manuscript.
